# Genetic liability to endometriosis and pregnancy outcomes: a two-sample Mendelian randomization study with maternal–fetal effect decomposition

**DOI:** 10.64898/2026.04.05.26350188

**Authors:** Jonas Vibert, Tuck Seng Cheng, Maria Christine Magnus, Elisabeth Aiton, Zoltán Kutalik, David Baud, Deborah A. Lawlor, Maria Carolina Borges, Nicola Pluchino

## Abstract

**Background:** Endometriosis is associated with adverse pregnancy outcomes in standard observational studies, including placental complications, preterm birth, and caesarean delivery. However, causal inference from these studies is complicated by residual confounding, differential clinical management, and the presence of intermediate factors such as subfertility and the use of assisted reproductive technologies, which may lie on the causal pathway between endometriosis and adverse outcomes.We applied Mendelian randomization (MR) to estimate the causal effects of genetic liability to endometriosis on a broad range of maternal and perinatal outcomes.

**Methods:** We conducted a two-sample MR study using summary-level GWAS data. Forty-one independent genetic instruments for endometriosis were derived from the largest available GWAS meta-analysis (60,674 cases; 701,926 controls; mean F-statistic = 279). SNP–outcome associations were obtained for 30 outcomes from the MR-PREG collaboration, FinnGen Release 12, and a postpartum haemorrhage GWAS meta-analysis, spanning placental disorders, pregnancy timing, labour and delivery, hypertensive disorders, fetal growth, and neonatal outcomes. Primary analyses used the inverse-variance weighted method, complemented by MR-Egger, weighted median, weighted mode, and MR-PRESSO. Trio-based models disentangled maternal from fetal genetic contributions. Multiple testing was addressed using false discovery rate correction.

**Findings:** Across 30 outcomes, only placenta praevia reached FDR-corrected significance, with a robust and consistent causal signal across four of five sensitivity methods (IVW OR 1·62, 95% CI 1·33–1·97; q<0·001). Within the placental disorders domain, estimates for premature placental separation and the broader placental disorders phenotype were directionally concordant but imprecise. For premature rupture of membranes, estimates were concordant across three methods, though the association was sensitive to cohort exclusion and did not survive multiple testing correction and should be interpreted cautiously. By contrast, hypertensive disorders, gestational diabetes, postpartum haemorrhage, stillbirth, and most neonatal outcomes showed estimates consistently close to the null across all methods. Trio-based analyses suggested predominantly maternal genetic pathways for most outcomes; fetal genetic contributions were not significant after correction for multiple testing, with exploratory signals observed for birthweight-related outcomes requiring independent replication.

**Interpretation:** A robust causal signal for placenta praevia alongside directionally consistent estimates across the placental disorders domain, suggests that mechanisms related to abnormal implantation and placentation may constitute a major mechanism for how endometriosis liability influences pregnancy. These results suggest that previously reported associations with broader obstetric outcomes may partly reflect confounding or clinical management patterns, and support targeted surveillance for abnormal placentation rather than a generalised elevation of obstetric risk.

**Funding:** TSC, MCB, EA, and DAL are members of the MRC Integrative Epidemiology Unit at the University of Bristol (MC_UU_00032/5). MCM is supported by the Research Council of Norway through its Centres of Excellence funding scheme (project No 262700); and the research project “Endometriosis and adenomyosis throughout the life-course” (project No 351058).

**Research in context panel:** *Evidence before this study:* We searched PubMed and MEDLINE from inception to March 2025 using combinations of the terms “endometriosis” and key pregnancy outcomes (including placenta praevia, preterm birth, postpartum haemorrhage, caesarean delivery, fetal growth, and pregnancy complications), without language restrictions. We included observational studies, systematic reviews, meta-analyses, and Mendelian randomisation studies. Observational evidence consistently suggests that women with endometriosis have increased risks of placenta praevia, preterm birth, caesarean delivery, and impaired fetal growth, with reported effect sizes for placenta praevia typically ranging from two- to four-fold. However, findings for other outcomes, including postpartum haemorrhage and hypertensive disorders, are inconsistent. Taken together, these findings suggest that endometriosis may primarily affect early implantation and placentation processes. Rather than supporting a general intensification of obstetric surveillance, our results highlight the need to better characterise the underlying biological mechanisms of abnormal placental implantation, with the aim of identifying targets for prevention. Mendelian randomisation studies have attempted to address these limitations, but results remain inconsistent. Some studies report little evidence for causal effects on pregnancy outcomes, while others suggest associations restricted to severe disease or specific outcomes. These discrepancies likely reflect differences in statistical power—particularly for rare outcomes—and outcome definitions. Importantly, no previous study has jointly examined a broad spectrum of pregnancy outcomes while disentangling maternal, fetal, and paternal genetic effects.

*Added value of this study:* Using a two-sample Mendelian randomisation design across 30 pregnancy and perinatal outcomes, we provide robust genetic evidence that liability to endometriosis causally increases the risk of placenta praevia. This association was consistent across multiple sensitivity analyses and supported by the absence of pleiotropy and heterogeneity. Across the broader placental disorders domain, estimates were directionally concordant, suggesting that the causal signal extends beyond a single phenotype and reflects a shared underlying mechanism related to implantation and placentation. In contrast, we found little evidence supporting a causal role of endometriosis liability in hypertensive disorders, gestational diabetes, postpartum haemorrhage, fetal growth restriction, or most neonatal outcomes. Associations observed for preterm birth and caesarean delivery were not robust to sensitivity analyses and appeared driven by pleiotropy or cohort-specific effects. By incorporating trio-based analyses, we further show that associations are predominantly driven by maternal genetic pathways, supporting a uterine mechanism rather than fetal genetic effects.

*Implications of all the available evidence:* Taken together, the available evidence supports a more targeted approach to pregnancy care in women with endometriosis. The robust and specific association with placenta praevia—together with directionally consistent findings across the placental disorders domain—supports targeted surveillance of placental location rather than a generalised increase in obstetric monitoring. The absence of consistent causal effects for most other outcomes suggests that previously reported associations in observational studies may largely reflect residual confounding, differences in the population of women who achieve pregnancy (for example due to subfertility or the use of assisted reproductive technologies), and variations in clinical management (such as increased surveillance or intervention rates), rather than a direct biological effect of endometriosis itself. From a mechanistic perspective, these findings point to impaired implantation and early placentation as the principal pathway linking endometriosis to adverse pregnancy outcomes. Future research should focus on the biology of uterine receptivity and placentation, including the role of disease severity and co-existing adenomyosis, to identify opportunities for early intervention.

## 1. INTRODUCTION

Endometriosis is a chronic oestrogen-dependent disease characterized by the abnormal presence of endometrial-like tissue outside the uterine cavity, leading to inflammation, adhesions, pain, and subfertility [1]. It affects approximately 10% of women of reproductive age and up to 50% of infertile women [2,3]. Beyond infertility, some observational studies suggest associations between endometriosis and adverse pregnancy outcomes, including hypertensive disorders, placental disorders, preterm birth, caesarean delivery, postpartum hemorrhage, stillbirth and fetal growth restriction [4–13].

These associations are inconsistent across studies [14], with effect estimates varying substantially by outcome, population, and mode of conception. Causal inference is further complicated by residual confounding by socioeconomic status, health-care utilisation, and infertility-related characteristics; selection bias from the overrepresentation of women conceiving through assisted reproductive technologies [15]; and differential misclassification arising from substantial underdiagnosis of endometriosis. Collectively, these biases may lead to over- or underestimation of true effects.

Mendelian randomization (MR) uses genetic variants as instrumental variables to estimate the causal effect of genetically predicted exposures on outcomes, thereby reducing bias from confounding and reverse causation [16]. Because alleles are randomly assorted at conception, MR capitalises on this natural experiment to provide a quasi-experimental framework for causal inference. MR can, under certain assumptions, help address key limitations of conventional observational studies by examining whether genetic liability (or predisposition) to endometriosis influences the risk of adverse pregnancy outcomes. In doing so, it offers an additional line of evidence to clarify how endometriosis may influence pregnancy health.

We therefore used a two-sample MR design to investigate the causal effects of genetic liability to endometriosis on a broad range of adverse pregnancy outcomes.

### 2. METHOD

#### 2.1. Study design

##### 2.1.1. Overview

We conducted a two-sample MR study to investigate the potential causal relationship between genetic liability to endometriosis and adverse pregnancy outcomes [17]. The study design is summarised in Figure 1. Independent genome-wide significant variants for endometriosis were selected as instruments from the GWAS meta-analysis by Rahmioglu *et al.* after linkage disequilibrium clumping, yielding 41 SNPs [18]. SNP–outcome associations were obtained for 30 outcomes from the Mendelian Randomization in Pregnancy (MR-PREG) collaboration [19], FinnGen release 12 [20], and the postpartum haemorrhage GWAS by Westergaard et al. [21]. The primary analysis used inverse-variance weighted MR, complemented by pleiotropy-robust sensitivity methods including MR-Egger, weighted median and weighted mode estimators, MR-PRESSO, heterogeneity tests, and leave-one-out analyses. For outcomes available in MR-PREG, we additionally leveraged trio-based models to test whether our main estimates could be influenced by offspring or paternal genotypes. Multiple testing was addressed using false discovery rate (FDR) p-value < 0.05.

**Figure 1.**
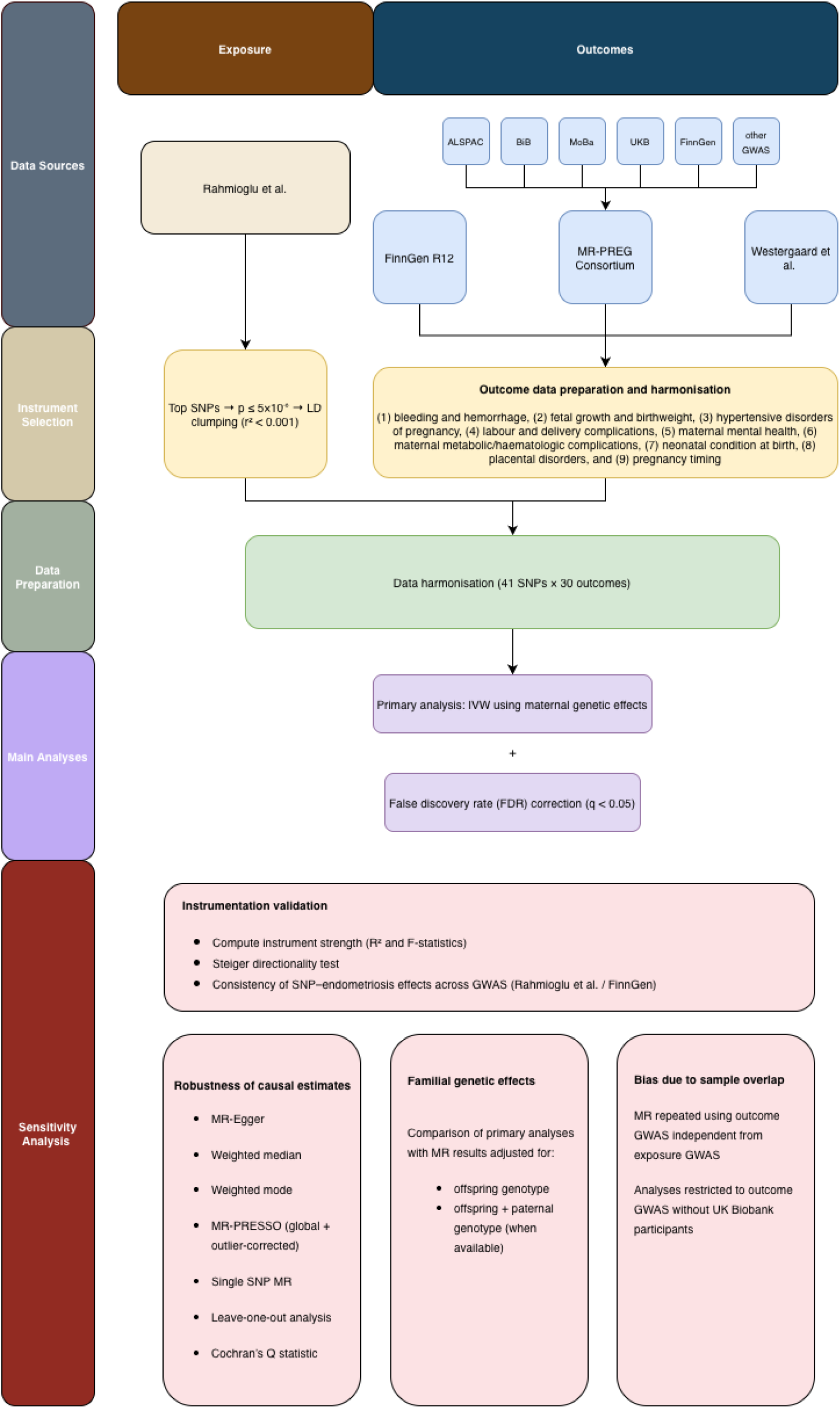
Study flowchart. *Genetic instruments for endometriosis were derived from a genome-wide association study (GWAS) by Rahmioglu et al., selecting genome-wide significant variants (p < 5 × 10*⁻⁸*) followed by linkage disequilibrium (LD) clumping (r² < 0.001). Outcome data were obtained from multiple large-scale GWAS consortia and cohorts, including MR-PREG (comprising ALSPAC, Born in Bradford [BiB], MoBa, UK Biobank, and additional GWAS), FinnGen (release R12), and Nordic registry-based studies*. *Outcome data were harmonised with genetic instruments, resulting in up to 41 independent variants across 30 pregnancy and perinatal outcomes. Primary analyses were conducted using inverse-variance weighted (IVW) Mendelian randomization based on maternal genetic effects, with false discovery rate (FDR) correction applied for multiple testing*. *Instrument validity was assessed using measures of instrument strength (R² and F-statistics), Steiger directionality tests, and consistency of SNP–endometriosis associations across GWAS. Sensitivity analyses included MR-Egger, weighted median and mode approaches, MR-PRESSO, single-SNP analyses, leave-one-out analyses, and heterogeneity testing (Cochran’s Q). Additional analyses evaluated familial genetic effects (including offspring and paternal genotypes where available) and potential bias due to sample overlap*.

#### 2.2. MR assumptions

For an MR instrument to be valid, it must satisfy three key assumptions: (i) *relevance*, whereby the genetic variants are strongly associated with endometriosis; (ii) *exchangeability*: there should be no confounding between genetic variants and outcomes in the population, which might be violated due to population stratification, assortative mating, or dynastic effects; and (iii) *exclusion restriction*, requiring that the variants influence adverse pregnancy outcomes exclusively through endometriosis liability. While only the first assumption can be empirically assessed (using F-statistics and variance explained), violations of independence and exclusion-restriction cannot be directly ruled out. Strategies used to assess or mitigate these violations are described below under ‘Sensitivity analyses’.

#### 2.3. Clinical grouping of outcomes

The 30 outcomes were grouped into nine clinical domains; full definitions and contributing cohorts are provided in Table 2 and Supplementary Table S1.

#### 2.4. Exposure data and instrument selection

##### 2.4.1. Exposure GWAS Source

Genetic variants were selected as instruments for endometriosis liability if they were associated with endometriosis at genome-wide significance (p < 5 × 10⁻⁸), using summary data from the largest available GWAS meta-analysis comprising approximately 60,674 cases and 701,926 controls of European ancestry (Rahmioglu et al., 2023 [18] (Table 1), after LD clumping (r² < 0.001 within a 10,000 kb window) using the European reference panel of the 1000 Genomes Project [22] (Supplementary Methods, Section 1.1 and 1.2).

**Table 1.** Exposure and outcome GWAS datasets included in the MR analyses.

| Study | Year | Dataset or consortium | Phenotype Group | Ancestry | Sample size | Main adjustments |
| --- | --- | --- | --- | --- | --- | --- |
| Rahmioglu et al. | 2023 | International Endometriosis Genetics Consortium + UK Biobank | Endometriosis (exposure) | Predominantly European (~98%) + Japanese (~2%) | 60,674 cases; 701,926 controls | Age, batch, principal components, study-specific covariates |
| McBride et al. (MR-PREG) | 2025 | MR-PREG collaboration (ALSPAC, BiB, MoBa, UKB, FinnGen + GWAS) | Adverse pregnancy and perinatal outcomes | Predominantly European | Up to 678,001 women | Age, study/centre, principal components |
| FinnGen R12 | 2024 | FinnGen (release 12) | Placental phenotypes | Finnish (European) | Up to 223,001 women | Age, batch, principal components |
| Westergaard et al. | 2024 | Nordic registry-based GWAS (6 cohorts) | Bleeding in pregnancy and postpartum hemorrhage | Northern European (registry-based) | Up to 331,792 women | Age, parity, calendar year, cohort |

Summary of genome-wide association studies (GWAS) used for the exposure (endometriosis) and pregnancy-related outcomes. For each dataset, the study source, year, phenotype group, ancestry, sample size, and main covariate adjustments are reported. The exposure GWAS from Rahmioglu et al. (2023) includes predominantly individuals of European ancestry with a small proportion of Japanese participants. Outcome GWAS were derived from large population-based consortia and cohorts, including MR-PREG, FinnGen, and Nordic registry-based studies. Sample sizes refer to the maximum number of women included in each GWAS. Adjustments correspond to covariates included in the original GWAS models.

A more recent multi-ancestry GWAS including over one million women has since been reported [23]; however, it was not available at the time of analysis. In addition, this GWAS incorporates multiple ancestries and includes data from direct-to-consumer cohorts, which may introduce heterogeneity in phenotype definition, and for which full summary statistics are not always publicly available. To ensure consistency with the predominantly European ancestry of the outcome datasets and to minimise potential bias due to population stratification, we restricted the primary analysis to instruments derived from a European-ancestry GWAS.

###### Instrument Selection

Instrument strength evaluation is detailed in Supplementary Methods, Section 1.3. Instrument strength was assessed using F-statistics derived from SNP–exposure associations; the mean per-SNP F-statistic was around 279, exceeding the conventional threshold of 10. However, given the large underlying GWAS sample size, these high F-statistics likely reflect statistical power rather than strong instrument–exposure associations. The instruments explained a moderate proportion of variance in the exposure (R² = 5.6%), indicating reasonable predictive strength. Methods for calculating R² and F-statistics, together with complete instrument characteristics, are described in Supplementary Methods, Section 1.3. Additional details on variant-level quality control, rsID mapping, LD pruning, harmonisation rules and SNP retention criteria are provided in Supplementary Methods Sections 1.1 and 1.2.

Although all 41 instruments passed harmonisation, the number of SNPs available per outcome ranged from 29 to 40 due to differences in SNP availability across outcome GWAS datasets. The exact SNP count per outcome is reported in Table 2.

**Table 2.**
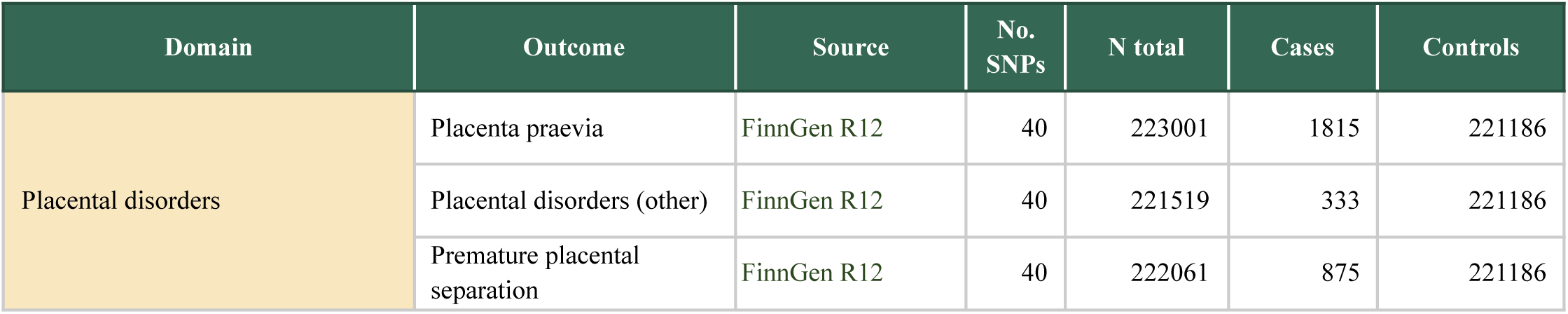
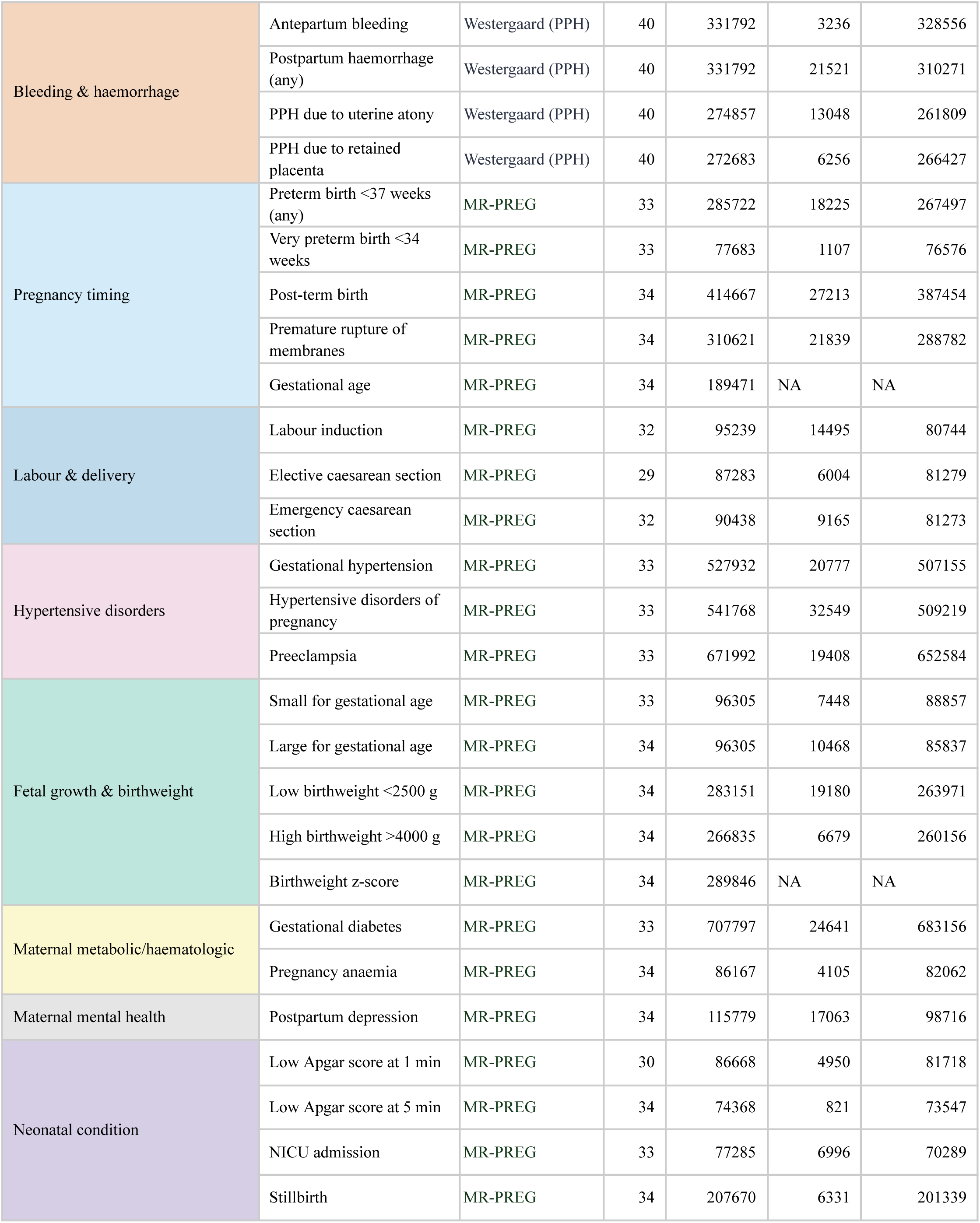
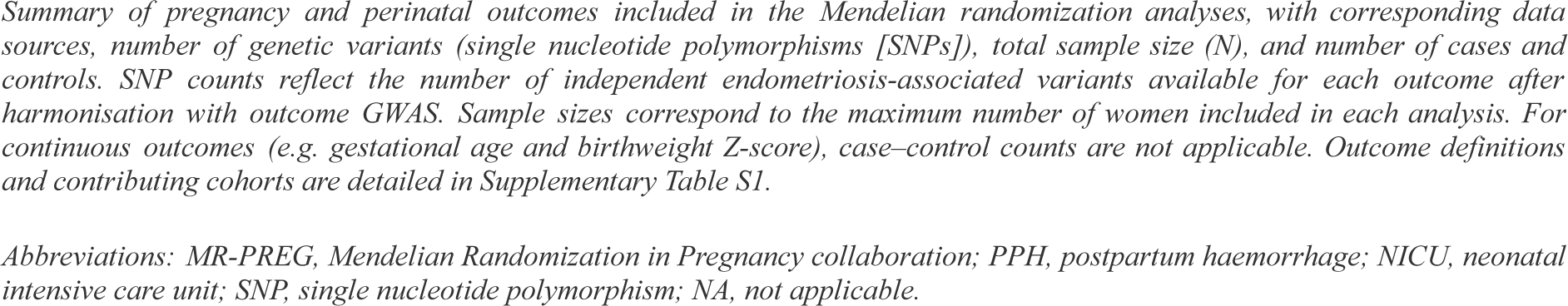
Outcome definitions, sample sizes, case/control counts, and SNP availability per outcome.

| Domain | Outcome | Source | No. SNPs | N total | Cases | Controls |
| --- | --- | --- | --- | --- | --- | --- |
| Placental disorders | Placenta praevia | FinnGen R12 | 40 | 223001 | 1815 | 221186 |
|  | Placental disorders (other) | FinnGen R12 | 40 | 221519 | 333 | 221186 |
|  | Premature placental separation | FinnGen R12 | 40 | 222061 | 875 | 221186 |
| Bleeding & haemorrhage | Antepartum bleeding | Westergaard (PPH) | 40 | 331792 | 3236 | 328556 |
|  | Postpartum haemorrhage (any) | Westergaard (PPH) | 40 | 331792 | 21521 | 310271 |
|  | PPH due to uterine atony | Westergaard (PPH) | 40 | 274857 | 13048 | 261809 |
|  | PPH due to retained placenta | Westergaard (PPH) | 40 | 272683 | 6256 | 266427 |
| Pregnancy timing | Preterm birth <37 weeks (any) | MR-PREG | 33 | 285722 | 18225 | 267497 |
|  | Very preterm birth <34 weeks | MR-PREG | 33 | 77683 | 1107 | 76576 |
|  | Post-term birth | MR-PREG | 34 | 414667 | 27213 | 387454 |
|  | Premature rupture of membranes | MR-PREG | 34 | 310621 | 21839 | 288782 |
|  | Gestational age | MR-PREG | 34 | 189471 | NA | NA |
| Labour & delivery | Labour induction | MR-PREG | 32 | 95239 | 14495 | 80744 |
|  | Elective caesarean section | MR-PREG | 29 | 87283 | 6004 | 81279 |
|  | Emergency caesarean section | MR-PREG | 32 | 90438 | 9165 | 81273 |
| Hypertensive disorders | Gestational hypertension | MR-PREG | 33 | 527932 | 20777 | 507155 |
|  | Hypertensive disorders of pregnancy | MR-PREG | 33 | 541768 | 32549 | 509219 |
|  | Preeclampsia | MR-PREG | 33 | 671992 | 19408 | 652584 |
| Fetal growth & birthweight | Small for gestational age | MR-PREG | 33 | 96305 | 7448 | 88857 |
|  | Large for gestational age | MR-PREG | 34 | 96305 | 10468 | 85837 |
|  | Low birthweight <2500 g | MR-PREG | 34 | 283151 | 19180 | 263971 |
|  | High birthweight >4000 g | MR-PREG | 34 | 266835 | 6679 | 260156 |
|  | Birthweight z-score | MR-PREG | 34 | 289846 | NA | NA |
| Maternal metabolic/haematologic | Gestational diabetes | MR-PREG | 33 | 707797 | 24641 | 683156 |
|  | Pregnancy anaemia | MR-PREG | 34 | 86167 | 4105 | 82062 |
| Maternal mental health | Postpartum depression | MR-PREG | 34 | 115779 | 17063 | 98716 |
| Neonatal condition | Low Apgar score at 1 min | MR-PREG | 30 | 86668 | 4950 | 81718 |
|  | Low Apgar score at 5 min | MR-PREG | 34 | 74368 | 821 | 73547 |
|  | NICU admission | MR-PREG | 33 | 77285 | 6996 | 70289 |
|  | Stillbirth | MR-PREG | 34 | 207670 | 6331 | 201339 |

Abbreviations: MR-PREG, Mendelian Randomization in Pregnancy collaboration; PPH, postpartum haemorrhage; NICU, neonatal intensive care unit; SNP, single nucleotide polymorphism; NA, not applicable.

###### Interpretation of estimates

Because endometriosis is a binary diagnosis arising from an underlying continuous liability, MR estimates should be interpreted as the effect of genetically predicted liability rather than as direct risk differences between women with and without clinically diagnosed disease. This distinction is a general limitation of MR analyses applied to categorical exposures and is particularly relevant when comparing estimates to conventional epidemiologic measures [24] (Supplementary Methods, Section 1.4).

##### 2.4.2. Outcome data

###### 2.4.2.1. MR-PREG

Summary-level GWAS data for adverse pregnancy outcomes were primarily obtained from the Mendelian Randomization in Pregnancy (MR-PREG) collaboration [19], which provides GWAS summary statistics across up to 714,972 women from multiple European cohorts for 34 maternal and fetal outcomes. The 23 outcomes examined in this study were identified *a priori* by clinicians (JV, DB and NP) on the basis of their biological and clinical plausibility of being influenced by endometriosis. The studies contributing to MR-PREG include ALSPAC, Born in Bradford, MoBa, UK Biobank, FinnGen, and publicly available GWAS. Detailed harmonisation procedures and cohort-specific information are provided in Supplementary Methods, Section 2.1, with phenotype definitions in Supplementary Table S1.

For outcomes not covered by MR-PREG, we used FinnGen Release 12 [20] and GWAS meta-analysis by Westergaard *et al.* [21].

###### 2.4.2.2. FinnGen (Placental disorders)

For placental disorders, we used FinnGen Release 12 [20], comprising up to 223,001 participants of predominantly Finnish ancestry. Case/control numbers for each phenotype are provided in Table 1, and further details are available in Supplementary Methods, Section 2.2.

###### 2.4.2.3. Westergaard et al. (Postpartum Hemorrhage)

For postpartum hemorrhage (PPH), we used the registry-based GWAS meta-analysis by Westergaard *et al.* [21], which included 21,521 cases from six Northern European cohorts of European ancestry, with subtype-specific analyses for uterine atony and retained placental tissue (Supplementary Methods, Section 2.3). Partial sample overlap is possible between the exposure GWAS and some outcome datasets, particularly UK Biobank and FinnGen (Supplementary Methods, Section 4). Despite the moderate variance explained, the high F-statistics (mean F = 279) indicate a low risk of weak instrument bias, and any bias arising from sample overlap is expected to be minimal.

###### 2.4.2.4. Maternal–Fetal Genotype Correction

Because several pregnancy outcomes may be influenced by both maternal and fetal genetic backgrounds [25,26], we used mutually adjusted effect estimates from the MR-PREG trio-based GWAS [20] obtained using a weighted linear model that jointly models maternal, fetal and paternal genotypes (Supplementary Methods, Section 6). This approach provides three conditioned estimates: the maternal genetic effect adjusted for fetal genotype (primary analysis), the fetal genetic effect adjusted for maternal genotype (sensitivity analysis), and the paternal effect used as a negative control. Without adjustment for fetal genotype, MR estimates based on maternal variants may be biased by fetal genetic confounding, whereby maternally transmitted alleles influence pregnancy outcomes through fetal or placental biology rather than maternal pathways, potentially violating the exclusion restriction assumption.

#### 2.5. Harmonisation

Effect alleles were harmonised across exposure and outcome datasets using the default harmonisation procedure implemented in TwoSampleMR. All palindromic SNPs (n=7) were retained since strand orientation could be reliably inferred from allele frequency information. Full harmonisation procedures are detailed in Supplementary Methods, Section 3.1.

#### 2.6. Statistical Analyses

##### 2.6.1. Primary MR Analysis

The primary MR analysis used the inverse-variance weighted (IVW) method [27] (see Supplementary Methods, Section 4.1). Effect estimates for binary outcomes are reported as odds ratios (ORs) with 95% confidence intervals (CIs), expressed per 1 unit increase in the log-odds of genetically predicted liability to endometriosis. Continuous outcomes are presented as β coefficients with 95% CIs on the same scale, expressed in weeks for gestational age and in standard deviation units for birthweight z-score.

We controlled for multiple testing using the false discovery rate (FDR), applying the Benjamini–Hochberg procedure to the IVW P-values. Associations with q < 0.05 were considered statistically significant (Supplementary Methods, Section 7). Because several outcomes are biologically and statistically correlated, interpretation did not rely solely on statistical significance; the magnitude, direction, and precision of estimates were also considered (Supplementary Methods, Section 5.7).

##### 2.6.2. Sensitivity Analyses

To mitigate bias from potential violations of core assumptions, we applied multiple sensitivity analyses including MR-Egger regression [28], the weighted median estimator [29], simple mode [30], weighted mode approaches [30], and MR-PRESSO [31] (Supplementary Methods, Sections 5.1–5.4). Directional horizontal pleiotropy was assessed through Egger intercept and PRESSO global tests. Heterogeneity across SNP-specific estimates was evaluated using Cochran’s Q test [17,32]. Single-SNP analyses were performed to examine the contribution of each instrument individually (18). Leave-one-out analyses were conducted at two levels: i) the SNP level, where each exposure SNP was sequentially excluded and ii) the cohort level, where each cohort included in the meta-analysis for each outcome was sequentially omitted to test the robustness of estimates to between-cohort heterogeneity (Supplementary Methods, Section 5.5). Consistency between IVW and sensitivity analyses was assessed based on concordance in effect direction and overlap of 95% confidence intervals. Because sensitivity estimators typically have lower statistical power than the IVW method, we considered results to be concordant across approaches, the directions of effect were consistent and the confidence intervals encompassed the IVW point estimate (Supplementary Methods, Sections 6).

#### 2.7. Software

We performed all analyses in R version 4.3.2 (R Foundation for Statistical Computing). Data preparation and harmonization were conducted using TwoSampleMR (version 0.5.6) and MRPRESSO (version 1.0.0). All analysis scripts and the full reproducible pipeline are publicly available in the endoMR-PREG GitHub repository (https://github.com/jonasvibert/endoMR-PREG). Further details regarding software implementation, package dependencies, and reproducibility procedures are provided in the Supplementary Methods, Section 8.

#### 2.9. Ethical Considerations

This study primarily used deidentified summary-level GWAS data and therefore did not require additional ethical approval. All original studies contributing to the GWAS of the outcomes of interest within the MR-PREG collaboration data obtained appropriate ethical approval and informed consent from participants. Further details are provided in Supplementary Methods, Section 9.

## 3. RESULTS

### Genetic instruments

From the GWAS of Rahmioglu et al., 9,259 SNPs were genome-wide significant for endometriosis (P < 5×10⁻⁸). After removing correlated variants based on LD, 41 independent SNPs were retained as genetic instruments (Supplementary Table S2). These variants collectively explained 5.6% of the variance in endometriosis liability, as estimated from the discovery GWAS summary statistics, and may therefore be slightly overestimated. The corresponding mean per-SNP F-statistic was 279, indicating a low risk of weak instrument bias.

Sample sizes, case and control counts, and the number of instruments available for each pregnancy outcome are shown in Table 2.

Across 30 outcomes, only one association reached FDR-corrected significance in the primary IVW analysis: placenta praevia (IVW OR 1·62, 95% CI 1·33–1·97; q<0·001). Results by domain are described below (Table 3, Figures 2–6).

**Figure 2.**
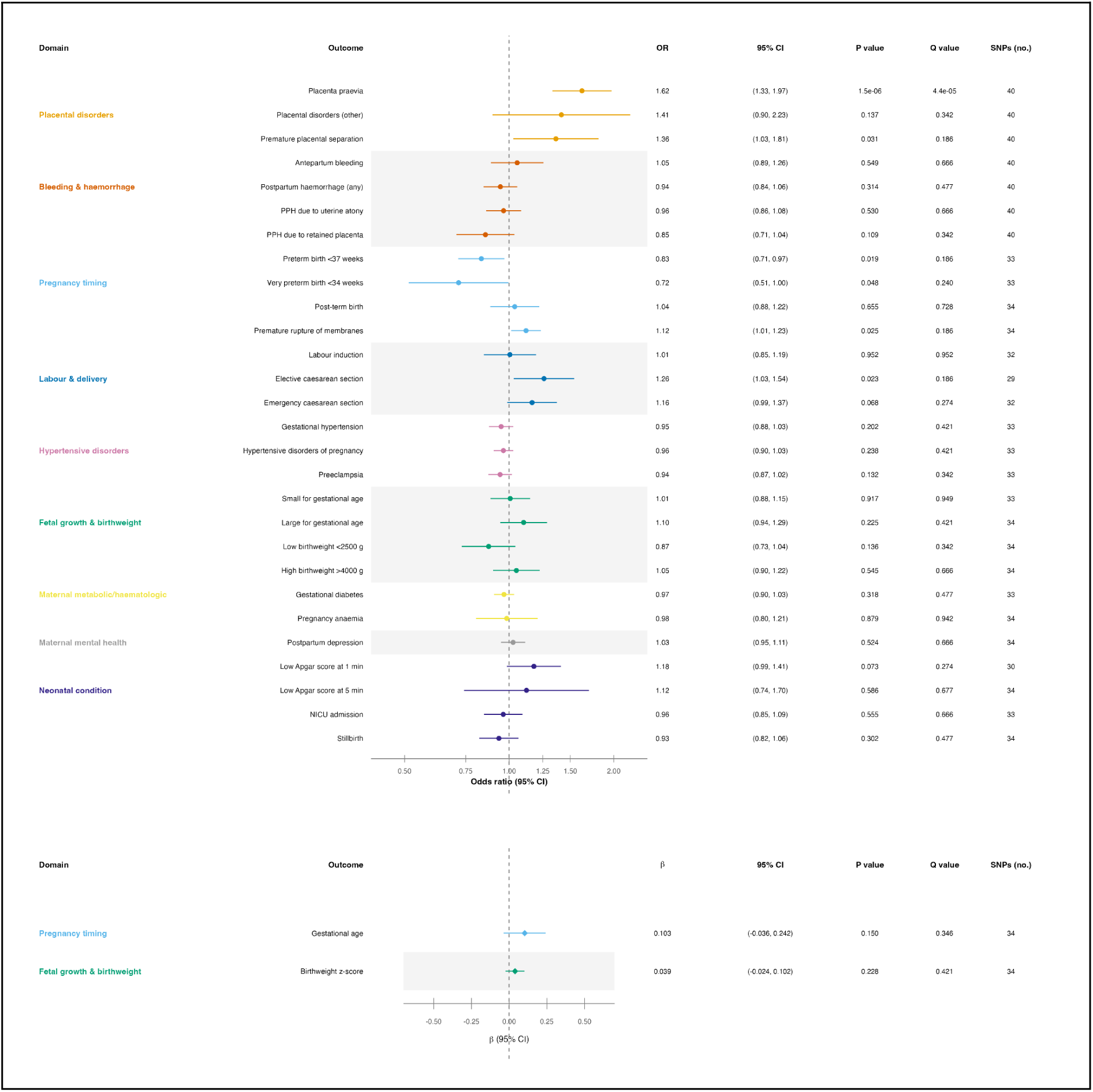
Inverse-variance weighted Mendelian randomization estimates for genetically predicted effects of endometriosis liability on pregnancy outcomes. Panel A: binary outcomes (reported as odds ratios). Panel B: continuous outcomes (reported as β coefficients per standard deviation increase in the outcome). *Forest plots showing inverse-variance weighted (IVW) Mendelian randomization estimates for the effect of genetically predicted liability to endometriosis on pregnancy and perinatal outcomes. Outcomes are grouped into clinical domains including placental disorders, bleeding and haemorrhage, pregnancy timing, labour and delivery, hypertensive disorders, fetal growth and birthweight, maternal metabolic or haematologic conditions, maternal mental health, and neonatal outcomes*. *In the upper panel, points represent odds ratios (ORs) for binary outcomes and horizontal lines indicate 95% confidence intervals. The vertical dashed line indicates the null value (OR = 1)*. *In the lower panel, points represent beta coefficients (β) for continuous outcomes (gestational age in weeks and birthweight z-score), with horizontal lines indicating 95% confidence intervals. The vertical dashed line indicates the null value (β = 0)*. *The number of SNP instruments used for each analysis is shown on the right, together with P values and false discovery rate–adjusted Q values. SNP counts vary across outcomes owing to differences in SNP availability across outcome GWAS datasets after harmonization*.

**Table 3.** IVW Mendelian randomization estimates for genetic liability to endometriosis and pregnancy outcomes (FDR-corrected)

| Domain | Outcome | Effect Scale | Estimate | 95% CI | P-value | q (FDR) | No. SNPs | Data source |
| --- | --- | --- | --- | --- | --- | --- | --- | --- |
| Placental disorders | Placenta praevia | OR | 1.62 | (1.33–1.97) | 1.46e-06 | 0.000 | 40 | FinnGen R12 |
|  | Placental disorders | OR | 1.41 | (0.90–2.23) | 0.14 | 0.343 | 40 | FinnGen R12 |
|  | Premature placental separation | OR | 1.36 | (1.03–1.81) | 0.03 | 0.186 | 40 | FinnGen R12 |
| Bleeding & haemorrhage | Antepartum bleeding | OR | 01.05 | (0.89–1.26) | 0.55 | 0.666 | 40 | Westergaard (PPH) |
|  | Postpartum hemorrhage (any) | OR | 0.94 | (0.84–1.06) | 0.31 | 0.477 | 40 | Westergaard (PPH) |
|  | PPH due to atony | OR | 0.96 | (0.86–1.08) | 0.53 | 0.666 | 40 | Westergaard (PPH) |
|  | PPH due to retained placenta | OR | 0.85 | (0.71–1.04) | 0.11 | 0.343 | 40 | Westergaard (PPH) |
| Pregnancy timing | Preterm birth <37 weeks (any) | OR | 0.83 | (0.71–0.97) | 0.02 | 0.186 | 33 | MR-PREG |
|  | Very preterm birth <34 weeks | OR | 0.72 | (0.51–1.00) | 0.05 | 0.240 | 33 | MR-PREG |
|  | Post-term birth | OR | 01.04 | (0.88–1.22) | 0.66 | 0.728 | 34 | MR-PREG |
|  | Premature rupture of membranes | OR | 1.12 | (1.01–1.23) | 0.03 | 0.186 | 34 | MR-PREG |
| | Gestational age | $\beta$ | 0.103 | (-0.036–0.242) | 0.15 | 0.346 | 34 | MR-PREG |
| Labour & delivery | Labour induction | OR | 01.01 | (0.85–1.19) | 0.95 | 0.952 | 32 | MR-PREG |
|  | Elective caesarean section | OR | 1.26 | (1.03–1.54) | 0.02 | 0.186 | 29 | MR-PREG |
|  | Emergency caesarean section | OR | 1.16 | (0.99–1.37) | 0.07 | 0.274 | 32 | MR-PREG |
| Hypertensive disorders | Gestational hypertension | OR | 0.95 | (0.88–1.03) | 0.20 | 0.420 | 33 | MR-PREG |
|  | Hypertensive disorders of pregnancy | OR | 0.96 | (0.90–1.03) | 0.24 | 0.420 | 33 | MR-PREG |
|  | Preeclampsia | OR | 0.94 | (0.87–1.02) | 0.13 | 0.343 | 33 | MR-PREG |
| Fetal growth & birthweight | Small for gestational age | OR | 01.01 | (0.88–1.15) | 0.92 | 0.949 | 33 | MR-PREG |
|  | Large for gestational age | OR | 1.10 | (0.94–1.29) | 0.23 | 0.420 | 34 | MR-PREG |
|  | Low birthweight <2500g | OR | 0.87 | (0.73–1.04) | 0.14 | 0.343 | 34 | MR-PREG |
|  | High birthweight >4000g | OR | 01.05 | (0.90–1.22) | 0.55 | 0.666 | 34 | MR-PREG |
| | Z-score birthweight | $\beta$ | 0.039 | (-0.024–0.102) | 0.23 | 0.420 | 34 | MR-PREG |
| Maternal metabolic/haematologic | Gestational diabetes | OR | 0.97 | (0.90–1.03) | 0.32 | 0.477 | 33 | MR-PREG |
|  | Pregnancy anemia | OR | 0.98 | (0.80–1.21) | 0.88 | 0.942 | 34 | MR-PREG |
| Maternal mental health | Postpartum depression | OR | 01.03 | (0.95–1.11) | 0.52 | 0.666 | 34 | MR-PREG |
| Neonatal condition | Low Apgar score at 1 min | OR | 1.18 | (0.99–1.41) | 0.07 | 0.274 | 30 | MR-PREG |
|  | Low Apgar score at 5 min | OR | 1.12 | (0.74–1.70) | 0.59 | 0.676 | 34 | MR-PREG |
|  | NICU admission | OR | 0.96 | (0.85–1.09) | 0.56 | 0.666 | 33 | MR-PREG |
|  | Stillbirth | OR | 0.93 | (0.82–1.06) | 0.30 | 0.477 | 34 | MR-PREG |

*Results are presented as odds ratios (OR) for binary outcomes and beta coefficients ( $\beta$ ) for continuous outcomes, with corresponding 95% confidence intervals (CI), p-values, and false discovery rate (FDR)-corrected q-values. Estimates were derived using inverse-variance weighted (IVW) Mendelian randomization. The number of genetic instruments (single nucleotide polymorphisms [SNPs]) reflects the number of independent endometriosis-associated variants available for each outcome after harmonisation.*
*FDR correction was applied across all outcomes to account for multiple testing. A q-value <0.05 was considered statistically significant.*
*Abbreviations: OR, odds ratio; CI, confidence interval; SNP, single nucleotide polymorphism; IVW, inverse-variance weighted; FDR, false discovery rate; NICU, neonatal intensive care unit; PPH, postpartum haemorrhage.*

### Placental disorders

Across all three placental outcomes, estimates were directionally consistent with an increased risk. The association of increased liability to endometriosis with placenta praevia was robust across four of five methods — IVW (OR 1·62, 95% CI 1·33–1·97), weighted median (OR 1·85, 95% CI 1·38–2·47), weighted mode (OR 2·09, 95% CI 1·23–3·54), and simple mode (OR 2·15, 95% CI 1·21–3·82) — with only MR-Egger yielding a wider, imprecise estimate, consistent with its lower power. For premature placental separation, IVW and weighted median were directionally concordant (OR 1·36, 95% CI 1·03–1·81 and OR 1·52, 95% CI 1·01–2·29), though the remaining methods did not converge which may reflect limited statistical power (875 cases). The broader placental disorders phenotype showed a positive point estimate across most methods but with substantial imprecision throughout (Figure 3 and Supplementary Figure S1).

**Figure 3.**
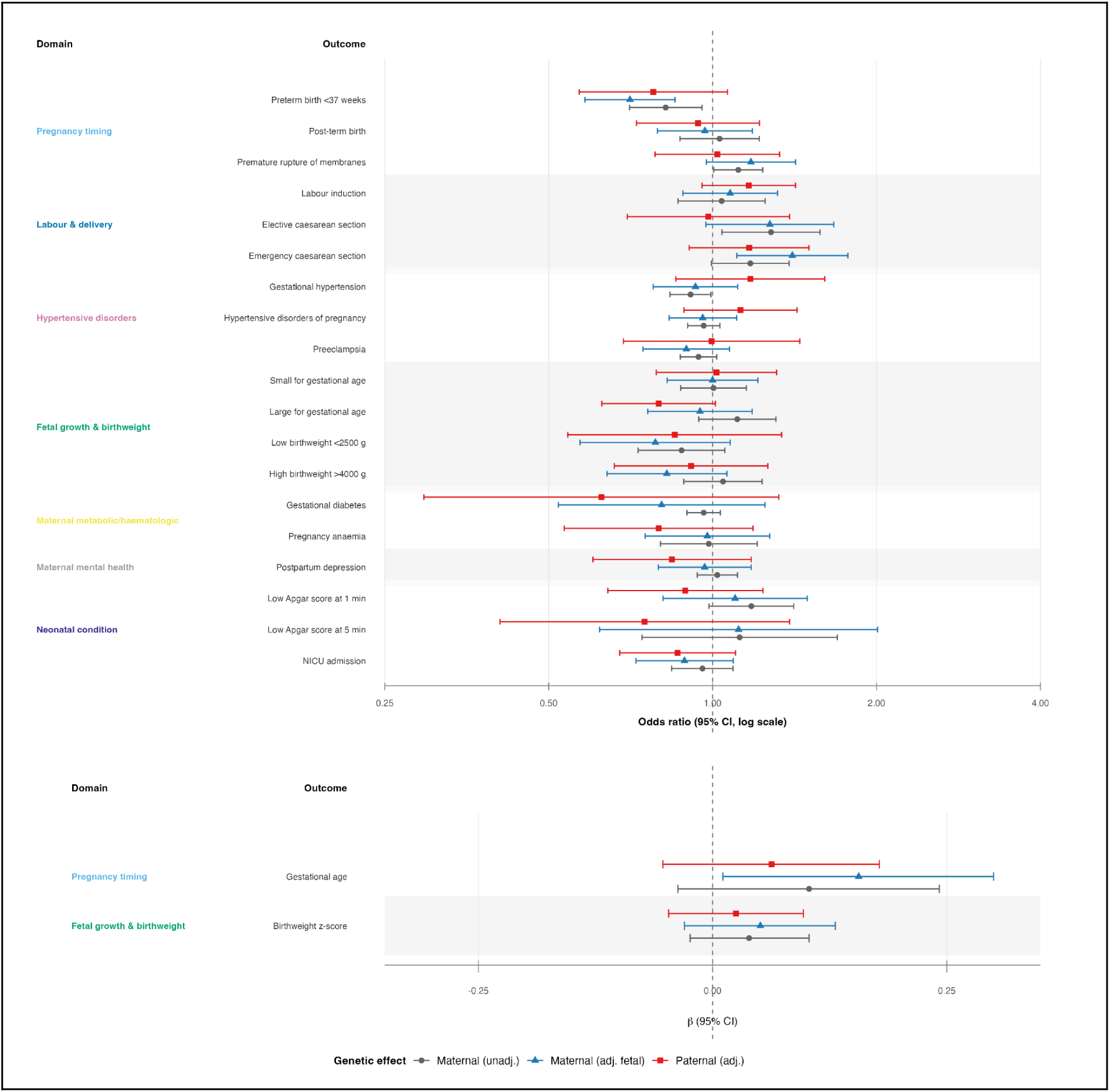
Trio-based decomposition of maternal, fetal, and paternal genetic effects of endometriosis liability on pregnancy outcomes. *Forest plots showing Mendelian randomization estimates for the effects of maternal, fetal, and paternal genetic liability to endometriosis on pregnancy and perinatal outcomes. Results are presented as odds ratios (OR) for binary outcomes (top panel) and beta coefficients (β) for continuous outcomes (bottom panel), with 95% confidence intervals. Estimates are shown for maternal effects without adjustment (unadjusted), maternal effects adjusted for fetal genotype, and paternal effects (where available). The vertical dashed line represents the null value (OR = 1 for binary outcomes; β = 0 for continuous outcomes). Outcomes are grouped by clinical domain*.

### Pregnancy timing

The IVW estimates suggested lower risk for preterm birth (OR 0·83, 95% CI 0·71–0·97) and very preterm birth (OR 0·72, 95% CI 0·51–1·00), but all four sensitivity methods were attenuated toward the null for both outcomes, with no method reaching nominal significance. Leave-one-cohort-out analyses showed both associations were driven by ALSPAC (Supplementary Table S5). This pattern of IVW-only signal with universal sensitivity method attenuation and single-cohort dependence suggests that these estimates may reflect horizontal pleiotropy or population-specific effects rather than a genuine causal association. Gestational age and post-term birth showed null IVW estimates, though the weighted median suggested modest positive associations for both — inconsistent with the primary analysis and not reproduced by other methods (Figure 3 and Supplementary Figure S2).

### Labour and delivery complications

Premature rupture of membranes showed convergent positive estimates across IVW (OR 1·12, 95% CI 1·01–1·23), weighted median (OR 1·14, 95% CI 1·03–1·28), and weighted mode (OR 1·23, 95% CI 1·02–1·47), with MR-Egger directionally consistent but imprecise. For elective caesarean section, the IVW was positive (OR 1·26, 95% CI 1·03–1·54) as was MR-Egger (OR 2·00, 95% CI 1·09–3·67), but weighted median, weighted mode, and simple mode were all close to unity, suggesting the signal may be driven by a subset of pleiotropic variants rather than reflecting a robust causal effect. Emergency caesarean and labour induction showed near null estimates across all methods (Figure 3 and Supplementary Figure S3).

### Neonatal condition at birth

Low Apgar score at 1 minute showed a positive IVW estimate (OR 1·18, 95% CI 0·99–1·41) reproduced by weighted median (OR 1·15, 95% CI 0·90–1·48) and MR-Egger, but weighted mode and simple mode were at unity, indicating at best a weak and uncertain signal. Low Apgar at 5 minutes showed positive point estimates across all methods but with uniformly wide confidence intervals, precluding firm conclusions. Neonatal intensive care unit (NICU) admission (IVW OR 0·96, 95% CI 0·85–1·09) and stillbirth (IVW OR 0·93, 95% CI 0·82–1·06) showed null estimates with no consistent directionality across methods (Figure 3 and Supplementary Figure S4).

### Bleeding, haemorrhage, hypertensive disorders, metabolic complications, and maternal mental health

Across these 10 outcomes, IVW estimates were consistently near unity and reproduced by weighted median, weighted mode, and simple mode estimates. MR-Egger yielded discordant estimates for PPH overall (OR 0·64, 95% CI 0·46–0·89) and PPH due to atony (OR 0·62, 95% CI 0·44–0·88), but these were inconsistent with all other methods and likely reflect horizontal pleiotropy and substantial heterogeneity rather than a true causal effect. These domains provide little evidence for a strong causal effect of endometriosis liability (Figure 3 and Supplementary Figure S5-8).

### Fetal growth and birthweight

Point estimates for LGA (IVW OR 1·10, 95% CI 0·94–1·29) and z-score birthweight (IVW β 0·04, 95% CI −0·02 to 0·10) were modestly positive, with weighted mode reaching nominal significance for birthweight z-score (β = 0.10, 95% CI 0.03 to 0.16, p = 0.008). Low birthweight showed a generally consistent negative direction across IVW, weighted median, and simple mode. While these patterns are directionally coherent with modestly larger offspring, effect sizes were small and no outcome reached FDR-corrected significance (Figure 3 and Supplementary Figure S9).

### Sensitivity analyses and robustness

Full results of heterogeneity and pleiotropy tests are provided in Supplementary Table S3, with domain-specific sensitivity plots in Supplementary Figures S1–S9. SNP-level leave-one-out analyses confirmed that no single variant drove the observed associations (Supplementary Figures S11–S16). Leave-one-cohort-out analyses showed that most associations were stable across datasets, with two exceptions: the association with very preterm birth was highly sensitive to exclusion of ALSPAC, and that with premature rupture of membranes attenuated substantially after its exclusion (Supplementary Table S5).

Overall, 15 of 30 outcomes (50%) showed full consistency across sensitivity methods and 7 (23%) were mostly consistent, defined as concordance in effect direction with the sensitivity method’s 95% CI encompassing the IVW point estimate (Supplementary Table S3). The association with placenta praevia showed no evidence of directional pleiotropy (Egger intercept P = 0·14), heterogeneity (Cochran’s Q P > 0·05), or outlier variants by MR-PRESSO. For preterm birth and very preterm birth, although formally classified as consistent by this definition, sensitivity method point estimates were substantially attenuated toward the null — and both associations were cohort-dependent — suggesting that formal consistency here reflects wide confidence intervals rather than convergent evidence. Only NICU admission was fully inconsistent across all sensitivity methods, against a null IVW estimate.

### Intergenerational (trio-based) analyses

Trio-based analyses were available for 21 outcomes (Supplementary Table S4; Figure 3). Overall, genetic liability to endometriosis was predominantly expressed through maternal genetic pathways, with paternal effects near the null for most outcomes. The strongest finding was for preterm birth, where the maternal effect adjusted for fetal genotype reached FDR significance (OR 0·71, 95% CI 0·58–0·85; q=0·007), though the paternal estimate was directionally concordant (OR 0·78; p=0·12), warranting caution. For emergency caesarean section, a nominally significant maternal effect (OR 1·40, 95% CI 1·11–1·77; p=0·005) was accompanied by an inverse fetal effect (OR 0·70; p=0·039), consistent with a maternal rather than fetal pathway. Exploratory fetal genetic signals were observed for LGA (OR 1·37, 95% CI 1·08–1·75; p=0·011) and high birthweight (OR 1·60, 95% CI 1·11–2·31; p=0·012), but did not survive multiple testing corrections and should be interpreted cautiously (Supplementary Figure S10).

## 4. DISCUSSION

In this two-sample MR analysis, genetic liability to endometriosis provided evidence consistent with a causal association with placenta praevia, supported across four of five sensitivity methods, with directionally concordant signals for premature placental separation and the broader placental disorders phenotype. For premature rupture of membranes, estimates were concordant across three methods, suggesting a plausible though more modest effect. By contrast, estimates for hypertensive disorders, gestational diabetes, postpartum haemorrhage, stillbirth, SGA, and most neonatal outcomes were precisely null across all methods, providing evidence against a causal role of endometriosis liability in these complications. For elective caesarean section and preterm birth, IVW signals were not reproduced by sensitivity methods, suggesting these reflect horizontal pleiotropy or clinical management patterns rather than independent causal effects.

Previous observational studies and meta-analyses report placenta praevia risks two to threefold higher in women with endometriosis, with even greater estimates in deep infiltrating disease (5–15). Our genetic findings support a causal interpretation: the endometriosis instruments predominantly capture moderate-to-severe disease pathways [18], consistent with the known disruption of eutopic endometrial function in these phenotypes. Mechanistically, impaired decidualisation and chronic low-grade inflammation in the eutopic endometrium may alter the implantation microenvironment, favouring low implantation sites and thereby increasing susceptibility to placenta praevia through altered implantation processes [34,35]. These findings suggest clinical relevance: endometriosis, particularly moderate to severe disease, may be considered an independent risk factor for placenta praevia, supporting careful assessment of placental location during routine ultrasound examinations, particularly in the presence of additional risk factors such as prior uterine surgery or assisted reproductive technologies.

The absence of robust causal signals for hypertensive disorders, gestational diabetes, and postpartum haemorrhage suggests that previously reported observational associations for these outcomes may largely reflect residual confounding, rather than endometriosis-specific biological pathways. Assisted reproductive technologies may also act as a potential mediating factor in these associations.. The inverse IVW association with preterm birth — not reproduced by any sensitivity method and entirely driven by a single cohort — most plausibly reflects cohort-specific ascertainment rather than a genuine protective effect. Accounting for fetal genetic effects add a further dimension: for most outcomes, findings were largely consistent with a predominant maternal genetic contribution, suggesting that effects are primarily mediated through the maternal environment rather than fetal genetic pathways. The notable exception was LGA and high birthweight, where fetal genetic contributions were more prominent, suggesting that endometriosis liability may influence fetal growth partly through fetal genetic pathways independent of maternal uterine environment.

Our findings substantially extend the existing Mendelian randomisation literature, which has thus far been limited in three key respects: a narrow range of outcomes that excluded important placental phenotypes, reliance on relatively underpowered GWAS sources for the endometriosis exposure, and the absence of triangulation of parental genetic effects [36–39]. The composite treatment of placenta praevia and placental abruption as a single outcome in prior work is particularly problematic, given that these conditions differ fundamentally in their underlying pathophysiology — one reflecting abnormal placental implantation, the other premature placental separation. By disaggregating these phenotypes, expanding the breadth of outcomes, and incorporating a maternal–fetal–paternal triangulation framework, we identify a robust and specific causal association with placenta praevia that, to our knowledge, has been insufficiently explored in previous MR studies.

Several limitations warrant consideration. Instruments predominantly capture liability to moderate-to-severe endometriosis (rASRM III–IV), reducing relevance for minimal or mild disease, and heterogeneity by subtype or anatomical phenotype could not be explored. Analyses were restricted to European-ancestry populations, limiting generalisability. Misclassification with adenomyosis remains possible and may differentially influence placentation-related outcomes. Genetic correlations between endometriosis and inflammatory or metabolic traits may represent residual sources of pleiotropy, particularly for hypertensive disorders and gestational diabetes. Finally, as MR estimates effects of genetic liability rather than clinical disease status, results should not be interpreted as risk differences between women with and without a diagnosis.

In conclusion, this MR study provides robust genetic evidence that liability to endometriosis causally increases the risk of placental complications, with placenta praevia showing the strongest and most consistent signal across methods, and directionally concordant estimates across the broader placental disorders domain. By contrast, estimates for hypertensive disorders, postpartum haemorrhage, fetal growth restriction, and most other obstetric outcomes were consistently near the null, suggesting that previously reported observational associations for these outcomes may largely reflect confounding, selection, or co-occurring conditions. Taken together, these findings suggest that endometriosis may primarily affect early implantation and placentation processes. Our results emphasise the need to better characterise the underlying biological mechanisms of abnormal placental implantation, with the aim of identifying targets for prevention.

## 5. CONTRIBUTORS

JV conceptualised the study, developed the methodology, curated and harmonised the data, conducted all formal analyses and sensitivity analyses, developed and maintained the analysis pipeline, generated the visualisations, and wrote the original draft. TSC contributed to methodology, including critical review and validation of the MR analysis pipeline and R code, and reviewed and edited the manuscript. MCM contributed resources and data curation for MoBa and reviewed and edited the manuscript. LA contributed resources and data curation for ALSPAC and BiB and reviewed and edited the manuscript. ZK contributed to methodology and statistical supervision and reviewed and edited the manuscript. DB contributed to conceptualisation, clinical framing, supervision, and manuscript review. DAL contributed to conceptualisation, co-establishment and scientific leadership of the MR-PREG collaboration, resources, funding acquisition, supervision, and manuscript review. MCB contributed to conceptualisation, methodology, resources, funding acquisition, supervision, and manuscript review, and directly accessed and verified the underlying data. NP contributed to conceptualisation, supervision, and manuscript review. JV and MCB directly accessed and verified all datasets used in the analyses. All authors interpreted the data, approved the final version of the manuscript, and accepted responsibility for the decision to submit for publication.

## Data Availability

Analytical R code is available on GitHub at: https://github.com/jonasvibert/endoMR-PREG
Genetic association data for adverse pregnancy outcomes generated by the MR-PREG collaboration can only be used for research that is covered by data agreements with current contributing studies.
The ALSPAC access policy that describes the proposal process in detail, including any costs associated with conducting research at ALSPAC, which may be updated from time to time, is available at: https://www.bristol.ac.uk/medialibrary/sites/alspac/documents/researchers/dataaccess/ALSPAC_Access_Policy.pdf
Data are available upon request from BiB, and information is available at: https://borninbradford.nhs.uk/research/how-to-access-data/
Data from MoBa are available upon application to Helsedata, administered by the Norwegian Institute of Public Health (see: https://helsedata.no for details).
Researchers can apply for access to UK Biobank data via the Access Management System (AMS): https://www.ukbiobank.ac.uk/enable-your-research/apply-for-access
Summary statistics from the endometriosis GWAS meta-analysis (Rahmioglu et al., 2023) excluding 23andMe are available from the EBI GWAS Catalog (Study Accession GCST90205183). GWAS summary statistics from 23andMe, Inc. were made available under a data use agreement that protects participant privacy; please contact or visit research.23andMe.com/collaborate for more information and to apply to access the data.
Meta-analysis summary statistics for PPH (Westergaard et al., 2024), including all subtypes, early bleeding, and antepartum haemorrhage, are deposited at: https://www.decode.com/summarydata
FinnGen Release 12 summary statistics are publicly available; full details are provided at: https://finngen.gitbook.io/documentation/

## 7. ACKNOWLEDGEMENTS

We are extremely grateful to all participants and families who took part in the contributing cohort studies, as well as the teams involved in data collection and management.

## AI disclosure statement

The authors used Claude (Anthropic, claude-sonnet-4-6) to assist with technical aspects of the analysis pipeline, including adapting existing R scripts to study-specific datasets, troubleshooting code, and generating visualisation scripts. ChatGPT (OpenAI, GPT-4) was used to improve the clarity and readability of the manuscript. No artificial intelligence tools were used to generate scientific hypotheses, interpret findings, or draw conclusions. All outputs were critically reviewed and validated by the authors, who take full responsibility for the content of the manuscript.

## 8. DATA SHARING STATEMENT

Analytical R code is available on GitHub at: https://github.com/jonasvibert/endoMR-PREG

Genetic association data for adverse pregnancy outcomes generated by the MR-PREG collaboration can only be used for research that is covered by data agreements with current contributing studies.

The ALSPAC access policy that describes the proposal process in detail, including any costs associated with conducting research at ALSPAC, which may be updated from time to time, is available at:

https://www.bristol.ac.uk/medialibrary/sites/alspac/documents/researchers/dataaccess/ALSPAC_Access_Policy.pdf

Data are available upon request from BiB, and information is available at: https://borninbradford.nhs.uk/research/how-to-access-data/

Data from MoBa are available upon application to Helsedata, administered by the Norwegian Institute of Public Health (see: https://helsedata.no for details).

Researchers can apply for access to UK Biobank data via the Access Management System (AMS): https://www.ukbiobank.ac.uk/enable-your-research/apply-for-access

Summary statistics from the endometriosis GWAS meta-analysis (Rahmioglu et al., 2023) excluding 23andMe are available from the EBI GWAS Catalog (Study Accession GCST90205183). GWAS summary statistics from 23andMe, Inc. were made available under a data use agreement that protects participant privacy; please contact or visit research.23andMe.com/collaborate for more information and to apply to access the data.

Meta-analysis summary statistics for PPH (Westergaard et al., 2024), including all subtypes, early bleeding, and antepartum haemorrhage, are deposited at: https://www.decode.com/summarydata

FinnGen Release 12 summary statistics are publicly available; full details are provided at: https://finngen.gitbook.io/documentation/

## TABLES AND FIGURES

### Tables (main manuscript)

- Table 1. Exposure and outcome GWAS datasets included in the MR analyses
- Table 2. Outcome definitions, sample sizes, case/control counts, and SNP availability per outcome
- Table 3. Primary IVW MR results across 30 outcomes (with FDR q-values)

### Figures (main manuscript)

- Figure 1. Study flowchart
- Figure 2. Overview forest plot of IVW estimates across outcomes
- Figure 3. Placental disorders domain (key/nominal signals)
- Figure 4. Pregnancy timing domain (key/nominal signals)
- Figure 5. Labour and delivery complications domain (key/nominal signals)
- Figure 6a and 6b. Trio-based analyses overview (maternal vs fetal vs paternal pathways)

### Supplementary Tables

- Supplementary Table S1. Detailed phenotype definitions and contributing cohorts (MR-PREG / FinnGen / Westergaard)
- Supplementary Table S2. Endometriosis instrument SNP list and characteristics (post-clumping)
- Supplementary Table S3. Sensitivity analyses including MR-Egger, weighted median, and weighted mode estimates, heterogeneity (Cochran’s Q), pleiotropy (Egger intercept), and MR-PRESSO results
- Supplementary Table S4. Trio-based MR estimates by maternal, fetal, and paternal genetic effects
- Supplementary Table S5. Leave-one-cohort-out sensitivity analysis: IVW estimates across all outcomes with each contributing study sequentially excluded

### Supplementary Figures

- Supplementary Figures S1–S9. Domain-specific Mendelian randomization results presented by outcome category.
- Supplementary Figure S10. Birthweight trio-based analysis (maternal vs fetal vs paternal effects)
- Supplementary Figure S11-16. SNP-level leave-one-out MR analyses

---

**Supplementary Table S1.**
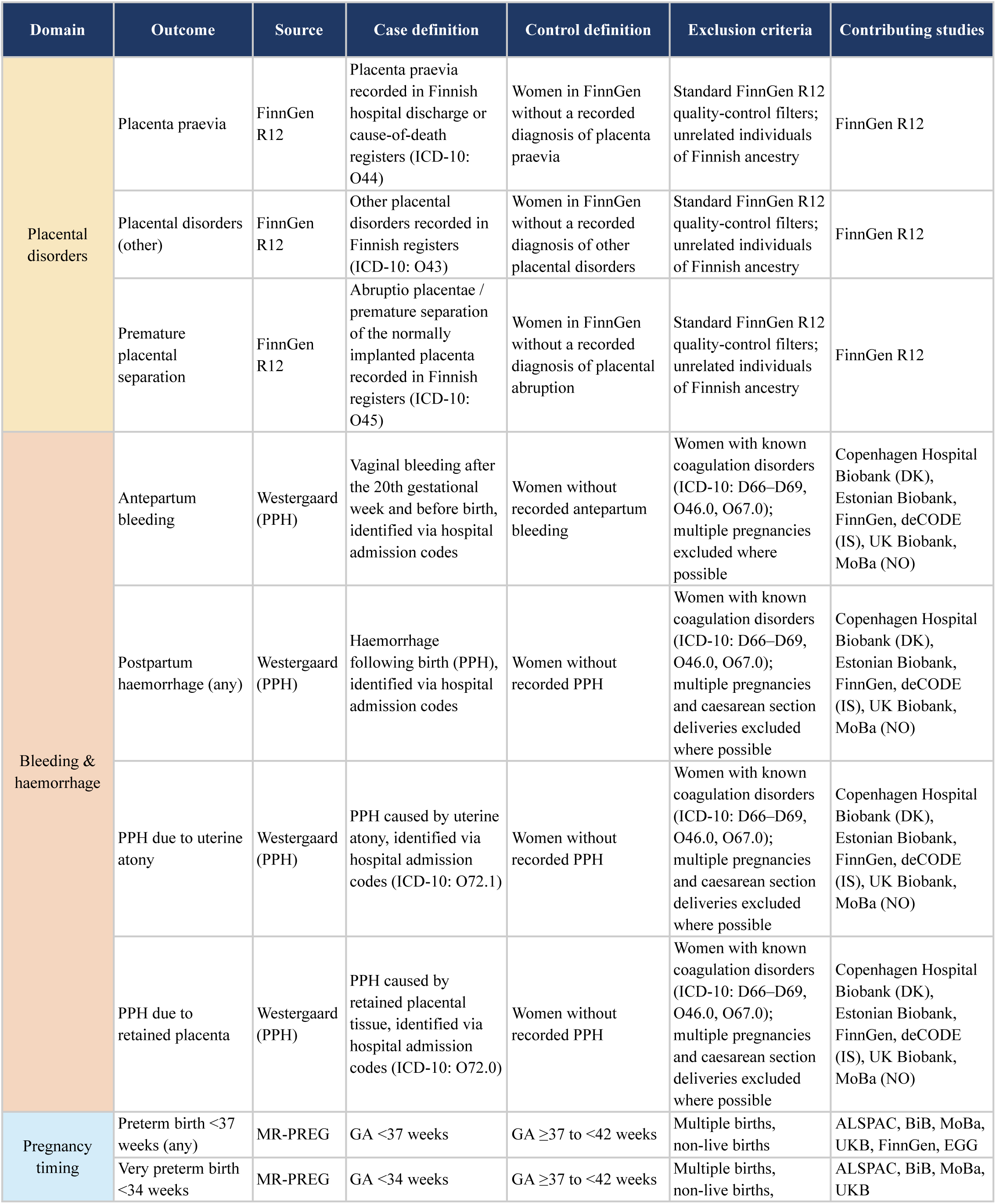
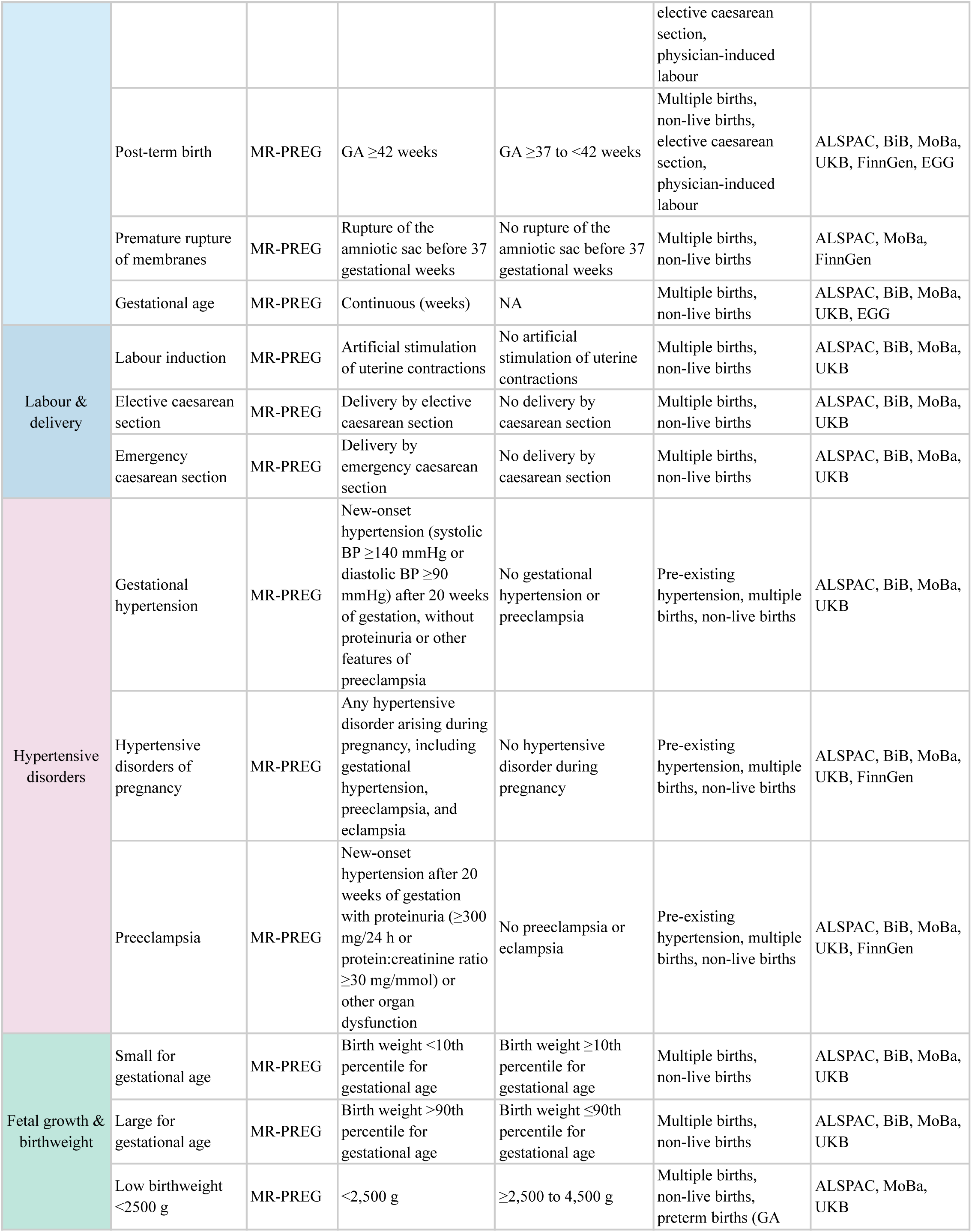
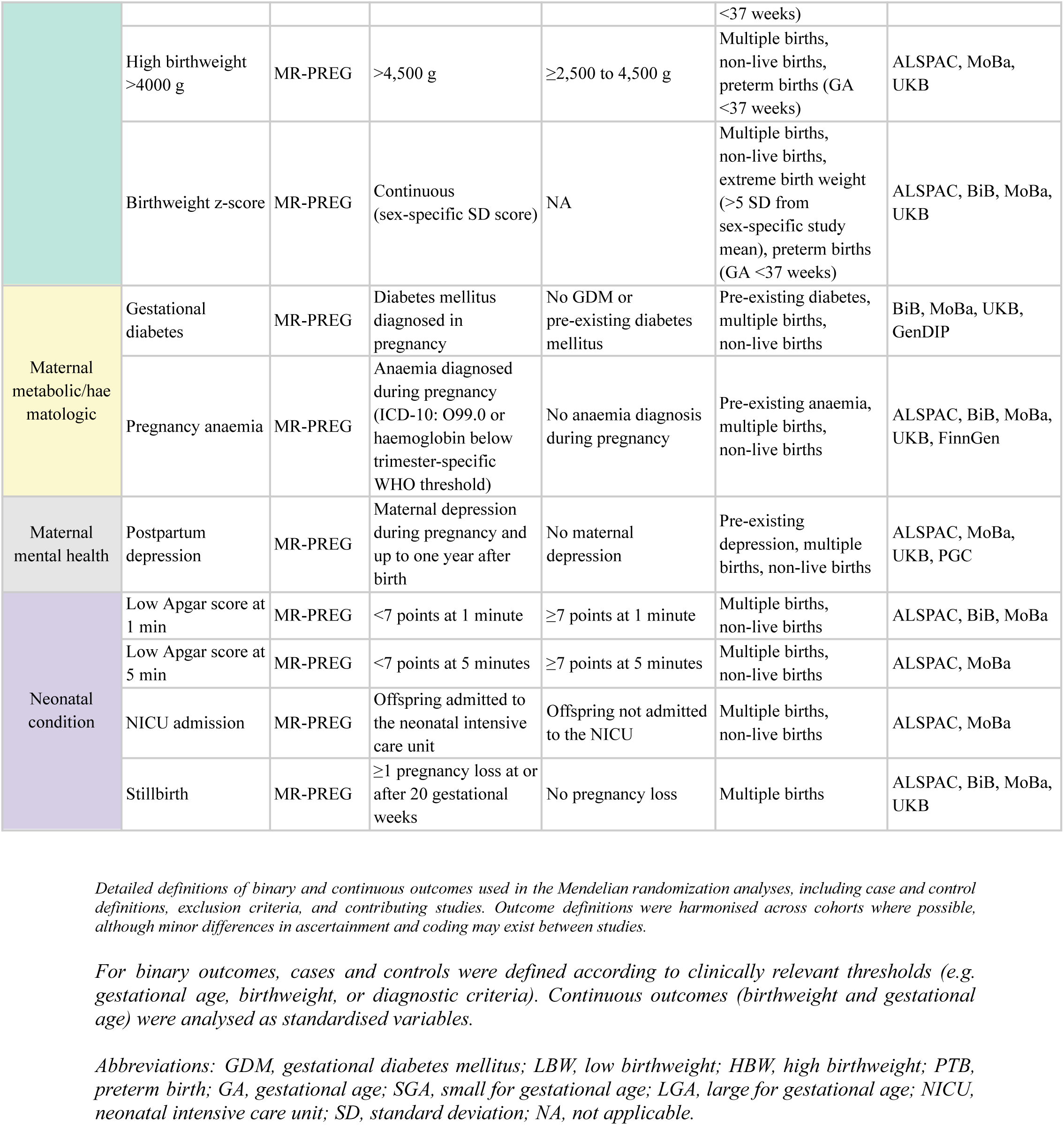
Definitions of pregnancy, maternal, labour and neonatal outcomes across contributing cohorts.

**Supplementary Table S2.** Characteristics of the genetic instruments for endometriosis (independent SNPs after clumping)

| SNP | Beta | SE | P-value | Effect allele | Other allele | EAF | r<sup>2</sup> | F |
| --- | --- | --- | --- | --- | --- | --- | --- | --- |
| rs12030576 | -0.0546 | 0.0075 | 3.34e-13 | T | G | 0.351 | 0.00136 | 280.04 |
| rs2421985 | -0.0532 | 0.0071 | 6.73e-14 | T | C | 0.5113 | 0.00141 | 291.64 |
| rs10917151 | 0.1322 | 0.0095 | 5.08e-44 | A | G | 0.1678 | 0.00488 | 1009.95 |
| rs10828249 | 0.0521 | 0.0075 | 3.74e-12 | A | G | 0.3531 | 0.00124 | 255.21 |
| rs7907732 | 0.0457 | 0.0072 | 2.19e-10 | T | C | 0.3867 | 0.000991 | 204.37 |
| rs3858429 | -0.1134 | 0.0099 | 2.23e-30 | T | C | 0.1567 | 0.0034 | 702.87 |
| rs7924571 | -0.052 | 0.0084 | 6.00e-10 | A | C | 0.2397 | 0.000986 | 203.33 |
| rs10860864 | -0.0564 | 0.009 | 3.69e-10 | T | C | 0.1838 | 0.000954 | 196.89 |
| rs56090796 | 0.0449 | 0.0076 | 3.46e-09 | A | T | 0.3255 | 0.000885 | 176.81 |
| rs3803042 | 0.0417 | 0.0072 | 6.97e-09 | A | G | 0.4235 | 0.000849 | 175.15 |
| rs12320196 | -0.0517 | 0.0069 | 6.74e-14 | A | G | 0.5268 | 0.00133 | 275.02 |
| rs7334326 | 0.0563 | 0.0099 | 1.29e-08 | C | G | 0.1569 | 0.000839 | 172.98 |
| rs57281976 | -0.0496 | 0.0084 | 3.53e-09 | A | G | 0.2342 | 0.000882 | 182.04 |
| rs12441483 | -0.0524 | 0.0071 | 1.58e-13 | T | C | 0.5283 | 0.00137 | 282.44 |
| rs66683298 | -0.0456 | 0.0074 | 7.18e-10 | T | C | 0.3974 | 0.000996 | 198.94 |
| rs7214750 | -0.0868 | 0.0142 | 9.80e-10 | T | C | 0.0698 | 0.000978 | 201.84 |
| rs2967684 | 0.0546 | 0.0096 | 1.29e-08 | A | G | 0.1577 | 0.000792 | 162.92 |
| rs11674184 | 0.0784 | 0.0074 | 3.16e-26 | T | G | 0.6084 | 0.00293 | 605.42 |
| rs6435157 | 0.0458 | 0.0084 | 4.97e-08 | T | C | 0.7654 | 0.000753 | 155.38 |
| rs1430787 | 0.0436 | 0.0076 | 9.65e-09 | A | T | 0.3052 | 0.000806 | 166.3 |
| rs1352889 | 0.0511 | 0.0092 | 2.79e-08 | T | C | 0.1734 | 0.000749 | 151.93 |
| rs1903068 | 0.0777 | 0.0076 | 1.55e-24 | A | G | 0.6787 | 0.00263 | 544.12 |
| rs2510770 | 0.0439 | 0.0076 | 7.64e-09 | A | G | 0.3211 | 0.00084 | 173.32 |
| rs2036754 | -0.0604 | 0.0109 | 3.00e-08 | A | T | 0.8785 | 0.000779 | 160.21 |
| rs2946160 | 0.0449 | 0.008 | 1.99e-08 | A | G | 0.738 | 0.00078 | 160.81 |
| rs2226158 | -0.0446 | 0.0071 | 3.35e-10 | A | G | 0.54 | 0.000988 | 203.88 |
| rs1971256 | -0.077 | 0.0085 | 1.32e-19 | T | C | 0.7891 | 0.00197 | 407.53 |
| rs73633307 | 0.1302 | 0.0192 | 1.19e-11 | T | G | 0.9574 | 0.00138 | 280.84 |
| rs71575922 | -0.1147 | 0.0098 | 1.21e-31 | C | G | 0.8472 | 0.00341 | 693.19 |
| rs11756073 | 0.0551 | 0.0093 | 3.13e-09 | A | G | 0.1711 | 0.000861 | 177.64 |
| rs6456259 | -0.0807 | 0.0096 | 4.23e-17 | A | G | 0.8398 | 0.00175 | 360.83 |
| rs4540228 | -0.0505 | 0.0075 | 1.66e-11 | T | C | 0.3362 | 0.00114 | 234.87 |
| rs1451383 | -0.0553 | 0.0081 | 8.66e-12 | A | G | 0.2567 | 0.00117 | 240.8 |
| rs55909142 | -0.0414 | 0.0072 | 8.92e-09 | T | C | 0.408 | 0.000828 | 170.79 |
| rs12549438 | -0.0534 | 0.0087 | 8.36e-10 | A | T | 0.7905 | 0.000944 | 188.29 |
| rs17053711 | -0.0459 | 0.008 | 9.61e-09 | A | G | 0.2694 | 0.000829 | 171.07 |
| rs10090060 | 0.0561 | 0.0072 | 6.61e-15 | A | G | 0.5719 | 0.00154 | 318.11 |
| rs10983311 | -0.047 | 0.0084 | 2.20e-08 | A | T | 0.2274 | 0.000776 | 159.8 |
| rs507666 | 0.0498 | 0.0087 | 1.04e-08 | A | G | 0.1951 | 0.000779 | 160.66 |
| rs10757277 | 0.0465 | 0.0071 | 5.78e-11 | A | G | 0.5157 | 0.00108 | 222.85 |
| rs10122243 | 0.0614 | 0.0072 | 1.49e-17 | T | C | 0.4051 | 0.00182 | 374.48 |

List of independent single nucleotide polymorphisms (SNPs) associated with endometriosis at genome-wide significance ( $p < 5 \times 10^{-8}$ ) and retained after linkage disequilibrium (LD) clumping. For each SNP, effect size (beta), standard error (SE), p-value, effect allele, other allele, effect allele frequency (EAF), proportion of variance explained ( $R^2$ ), and F-statistic are reported.
$R^2$ values represent the variance in endometriosis liability explained by each SNP. F-statistics were calculated to assess instrument strength, with values $>10$ indicating strong instruments. All SNPs were harmonised with outcome datasets prior to analysis.

**Supplementary Table S3.**
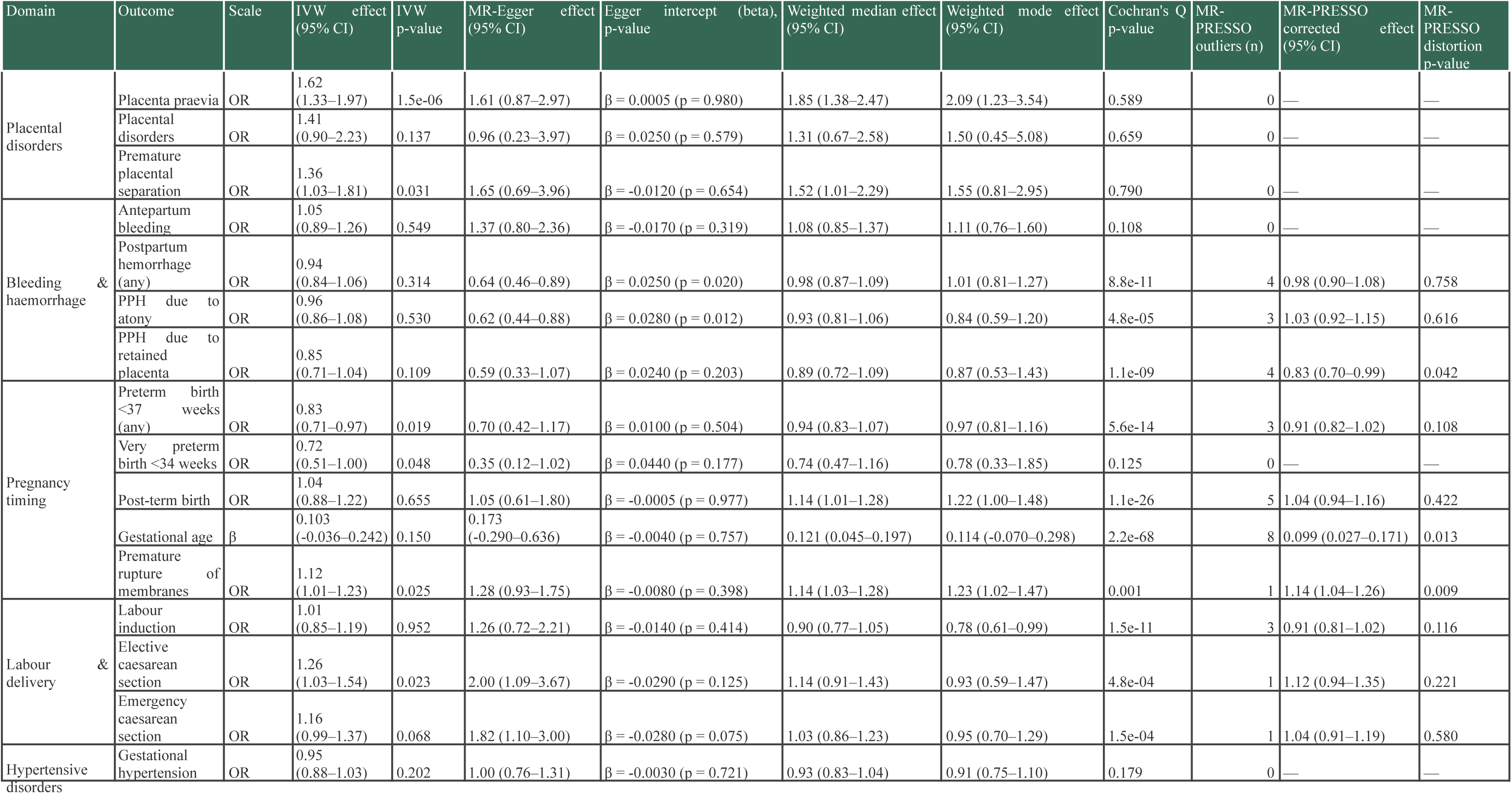
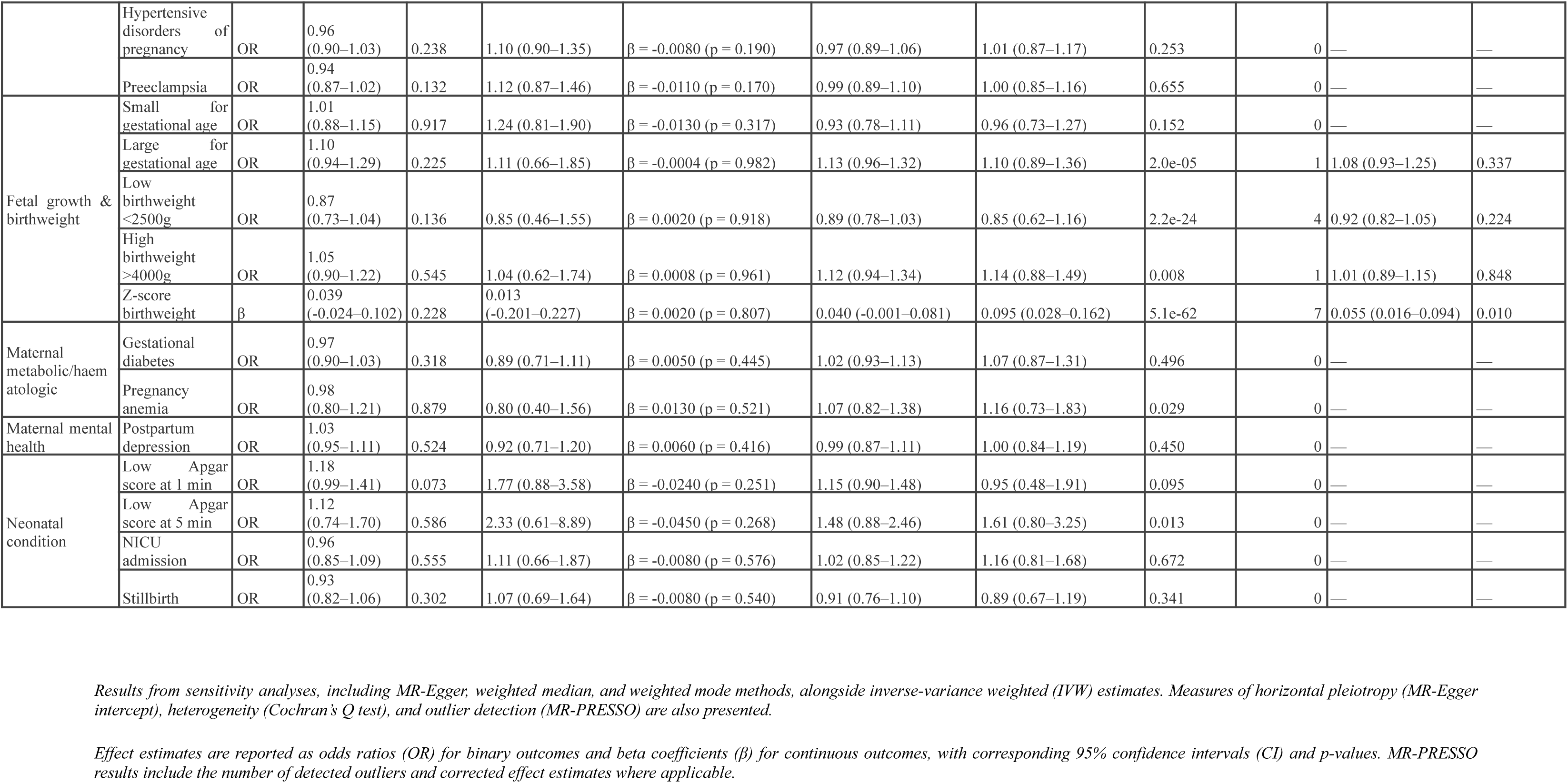
Sensitivity analyses including MR-Egger, weighted median, and weighted mode estimates, heterogeneity (Cochran’s Q), pleiotropy (Egger intercept), and MR-PRESSO results.

**Supplementary Table S4.** Trio-based Mendelian randomization estimates for the effect of genetic liability to endometriosis on pregnancy and perinatal outcomes by maternal, fetal, and paternal origin.

| Outcome | origin | nsnp | b | se | OR | LCL | UCL | pval | qval |
| --- | --- | --- | --- | --- | --- | --- | --- | --- | --- |
| Elective caesarean section | Maternal | 27 | 0.247 | 0.106 | 1.28 | 01.04 | 1.575 | 0.02 | 0.314 |
|  | Fetal | 27 | 0.124 | 0.122 | 1.132 | 0.892 | 1.437 | 0.309 | 0.764 |
|  | Paternal | 27 | -0.009 | 0.106 | 0.991 | 0.805 | 1.221 | 0.933 | 0.997 |
| Emergency caesarean section | Maternal | 30 | 0.159 | 0.084 | 1.173 | 0.995 | 1.382 | 0.0567 | 0.51 |
|  | Fetal | 30 | -0.112 | 0.094 | 0.894 | 0.743 | 1.075 | 0.234 | 0.703 |
|  | Paternal | 30 | -0.024 | 0.08 | 0.976 | 0.835 | 1.142 | 0.766 | 0.968 |
| Gestational age | Maternal | 34 | 0.103 | 0.071 | NA | NA | NA | 0.15 | 0.695 |
|  | Fetal | 34 | 0.001 | 0.048 | NA | NA | NA | 0.978 | 0.997 |
|  | Paternal | 34 | 0.01 | 0.039 | NA | NA | NA | 0.799 | 0.968 |
| Gestational diabetes | Maternal | 31 | -0.038 | 0.036 | 0.963 | 0.897 | 1.033 | 0.291 | 0.764 |
|  | Fetal | 31 | 0.016 | 0.23 | 1.016 | 0.647 | 1.596 | 0.944 | 0.997 |
|  | Paternal | 31 | -0.273 | 0.276 | 0.761 | 0.443 | 1.308 | 0.323 | 0.764 |
| Gestational hypertension | Maternal | 30 | -0.094 | 0.044 | 0.911 | 0.835 | 0.993 | 0.0335 | 0.42 |
|  | Fetal | 30 | -0.004 | 0.067 | 0.996 | 0.874 | 1.135 | 0.954 | 0.997 |
|  | Paternal | 30 | 0.136 | 0.102 | 1.146 | 0.938 | 1.4 | 0.183 | 0.695 |
| High birthweight >4000g | Maternal | 31 | 0.044 | 0.084 | 1.045 | 0.885 | 1.233 | 0.604 | 0.958 |
|  | Fetal | 31 | 0.332 | 0.106 | 1.394 | 1.132 | 1.715 | 0.00172 | 0.108 |
|  | Paternal | 31 | 0.147 | 0.112 | 1.158 | 0.929 | 1.443 | 0.192 | 0.695 |
| Hypertensive disorders of pregnancy | Maternal | 30 | -0.037 | 0.035 | 0.963 | 0.9 | 1.031 | 0.284 | 0.764 |
|  | Fetal | 30 | 0.052 | 0.058 | 1.053 | 0.94 | 1.179 | 0.37 | 0.765 |
|  | Paternal | 30 | 0.126 | 0.08 | 1.134 | 0.97 | 1.325 | 0.115 | 0.692 |
| Labour induction | Maternal | 30 | 0.038 | 0.094 | 1.039 | 0.864 | 1.249 | 0.684 | 0.958 |
|  | Fetal | 30 | 0.041 | 0.057 | 1.042 | 0.932 | 1.165 | 0.469 | 0.87 |
|  | Paternal | 30 | 0.118 | 0.068 | 1.125 | 0.985 | 1.285 | 0.0814 | 0.57 |
| Large for gestational age | Maternal | 32 | 0.104 | 0.083 | 1.11 | 0.943 | 1.307 | 0.21 | 0.695 |
|  | Fetal | 32 | 0.177 | 0.065 | 1.193 | 1.051 | 1.355 | 0.00641 | 0.202 |
|  | Paternal | 32 | -0.07 | 0.082 | 0.932 | 0.794 | 1.095 | 0.392 | 0.772 |
| Low Apgar score at 1 min | Maternal | 30 | 0.164 | 0.091 | 1.178 | 0.985 | 1.409 | 0.0729 | 0.57 |
|  | Fetal | 30 | 0.123 | 0.091 | 1.131 | 0.946 | 1.352 | 0.178 | 0.695 |
|  | Paternal | 30 | -0.047 | 0.108 | 0.954 | 0.773 | 1.178 | 0.66 | 0.958 |
| Low Apgar score at 5 min | Maternal | 34 | 0.115 | 0.211 | 1.121 | 0.742 | 1.695 | 0.586 | 0.958 |
|  | Fetal | 34 | -0.091 | 0.259 | 0.913 | 0.549 | 1.518 | 0.726 | 0.958 |
|  | Paternal | 34 | -0.283 | 0.221 | 0.753 | 0.489 | 1.161 | 0.199 | 0.695 |
| Low birthweight <2500g | Maternal | 33 | -0.132 | 0.094 | 0.877 | 0.73 | 1.053 | 0.159 | 0.695 |
|  | Fetal | 33 | 0.001 | 0.126 | 1.001 | 0.782 | 1.281 | 0.993 | 0.997 |
|  | Paternal | 33 | -0.063 | 0.144 | 0.939 | 0.709 | 1.245 | 0.663 | 0.958 |
| NICU admission | Maternal | 32 | -0.043 | 0.066 | 0.958 | 0.841 | 01.09 | 0.512 | 0.896 |
|  | Fetal | 32 | 0.019 | 0.08 | 1.019 | 0.871 | 1.192 | 0.815 | 0.968 |
|  | Paternal | 32 | -0.073 | 0.08 | 0.929 | 0.794 | 1.087 | 0.36 | 0.765 |
| Post-term birth | Maternal | 33 | 0.03 | 0.086 | 1.03 | 0.871 | 1.218 | 0.73 | 0.958 |
|  | Fetal | 33 | 0.059 | 0.071 | 1.061 | 0.922 | 1.22 | 0.409 | 0.782 |
|  | Paternal | 33 | 0 | 0.093 | 1 | 0.834 | 1.199 | 0.997 | 0.997 |
| Postpartum depression | Maternal | 32 | 0.02 | 0.043 | 1.02 | 0.937 | 1.111 | 0.647 | 0.958 |
|  | Fetal | 32 | 0.002 | 0.085 | 1.002 | 0.848 | 1.184 | 0.98 | 0.997 |
|  | Paternal | 32 | -0.121 | 0.124 | 0.886 | 0.695 | 1.129 | 0.328 | 0.764 |
| Preeclampsia | Maternal | 33 | -0.06 | 0.04 | 0.942 | 0.872 | 1.018 | 0.132 | 0.692 |
|  | Fetal | 33 | 0.048 | 0.072 | 1.049 | 0.91 | 1.209 | 0.51 | 0.896 |
|  | Paternal | 33 | 0.047 | 0.131 | 1.048 | 0.811 | 1.353 | 0.72 | 0.958 |
| Pregnancy anemia | Maternal | 34 | -0.016 | 0.104 | 0.984 | 0.802 | 1.208 | 0.879 | 0.997 |
|  | Fetal | 34 | -0.117 | 0.107 | 0.89 | 0.721 | 1.098 | 0.276 | 0.764 |
|  | Paternal | 34 | -0.209 | 0.139 | 0.811 | 0.618 | 1.065 | 0.132 | 0.692 |
| Premature rupture of membranes | Maternal | 32 | 0.109 | 0.053 | 1.115 | 1.005 | 1.236 | 0.04 | 0.42 |
|  | Fetal | 32 | -0.02 | 0.055 | 0.98 | 0.88 | 1.091 | 0.71 | 0.958 |
|  | Paternal | 32 | -0.038 | 0.08 | 0.963 | 0.824 | 1.126 | 0.638 | 0.958 |
| Preterm birth <37 weeks (any) | Maternal | 32 | -0.198 | 0.078 | 0.82 | 0.704 | 0.956 | 0.011 | 0.231 |
|  | Fetal | 32 | 0.001 | 0.084 | 1.001 | 0.848 | 1.181 | 0.993 | 0.997 |
|  | Paternal | 32 | -0.098 | 0.106 | 0.907 | 0.737 | 1.116 | 0.356 | 0.765 |
| Small for gestational age | Maternal | 30 | 0.004 | 0.071 | 1.004 | 0.874 | 1.153 | 0.957 | 0.997 |
|  | Fetal | 30 | 0.018 | 0.072 | 1.018 | 0.883 | 1.173 | 0.805 | 0.968 |
|  | Paternal | 30 | 0.021 | 0.088 | 1.021 | 0.86 | 1.212 | 0.812 | 0.968 |
| Z-score birthweight | Maternal | 34 | 0.039 | 0.032 | NA | NA | NA | 0.228 | 0.703 |
|  | Fetal | 34 | 0.015 | 0.017 | NA | NA | NA | 0.376 | 0.765 |
|  | Paternal | 34 | 0.013 | 0.022 | NA | NA | NA | 0.55 | 0.937 |

Results are presented separately for maternal, fetal, and paternal genetic effects. For each outcome, the number of SNPs included, effect estimate (b), standard error (SE), odds ratio (OR), 95% confidence interval (CI), p-value, and false discovery rate (FDR)-corrected q-value are shown. For continuous outcomes, odds ratios and confidence intervals are not applicable and are therefore reported as NA. Trio-based analyses were used to distinguish maternal genetic effects from fetal and paternal genetic contributions, helping to clarify whether observed associations may reflect intrauterine, inherited, or broader familial mechanisms.

**Supplementary Table S5.**
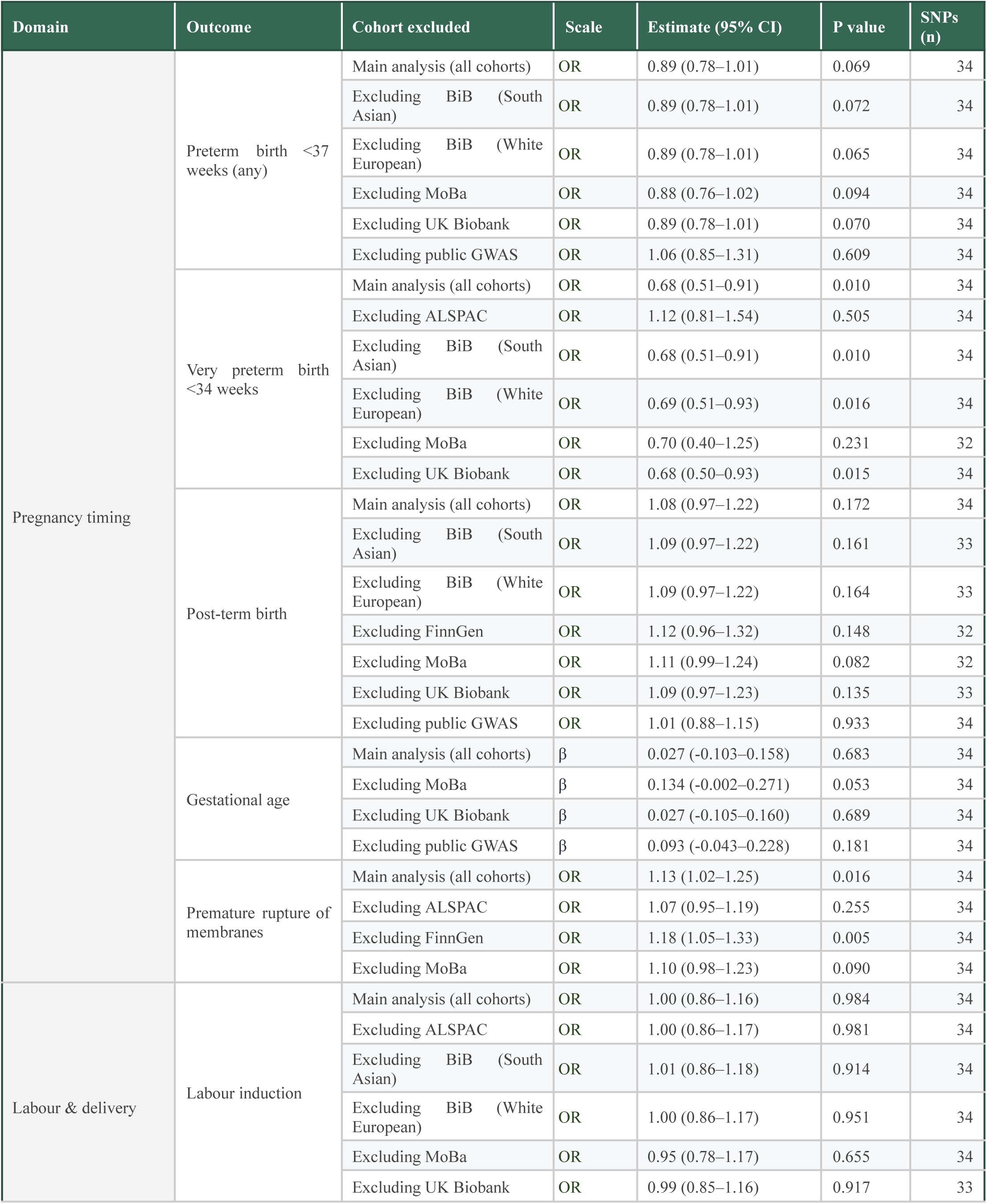
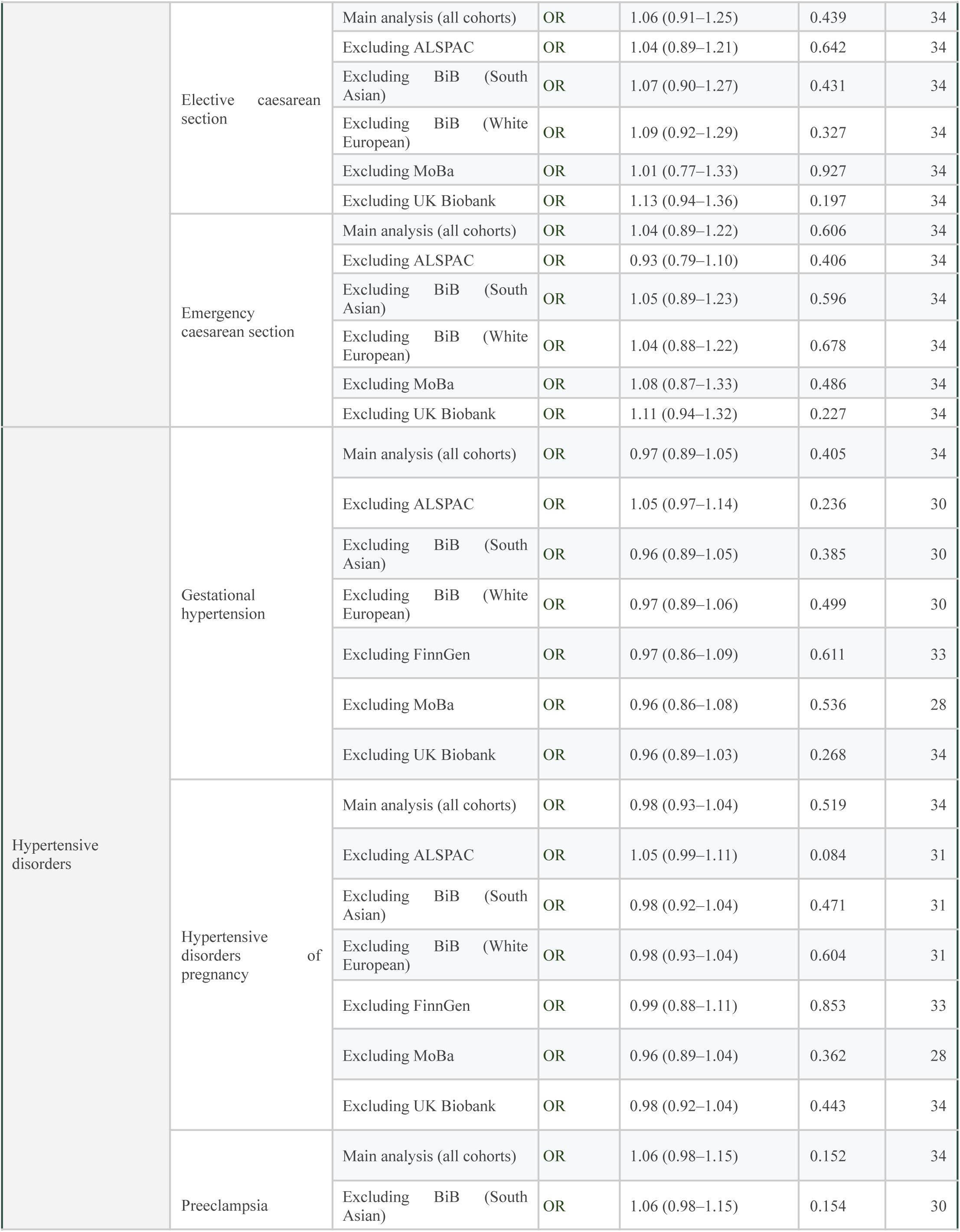
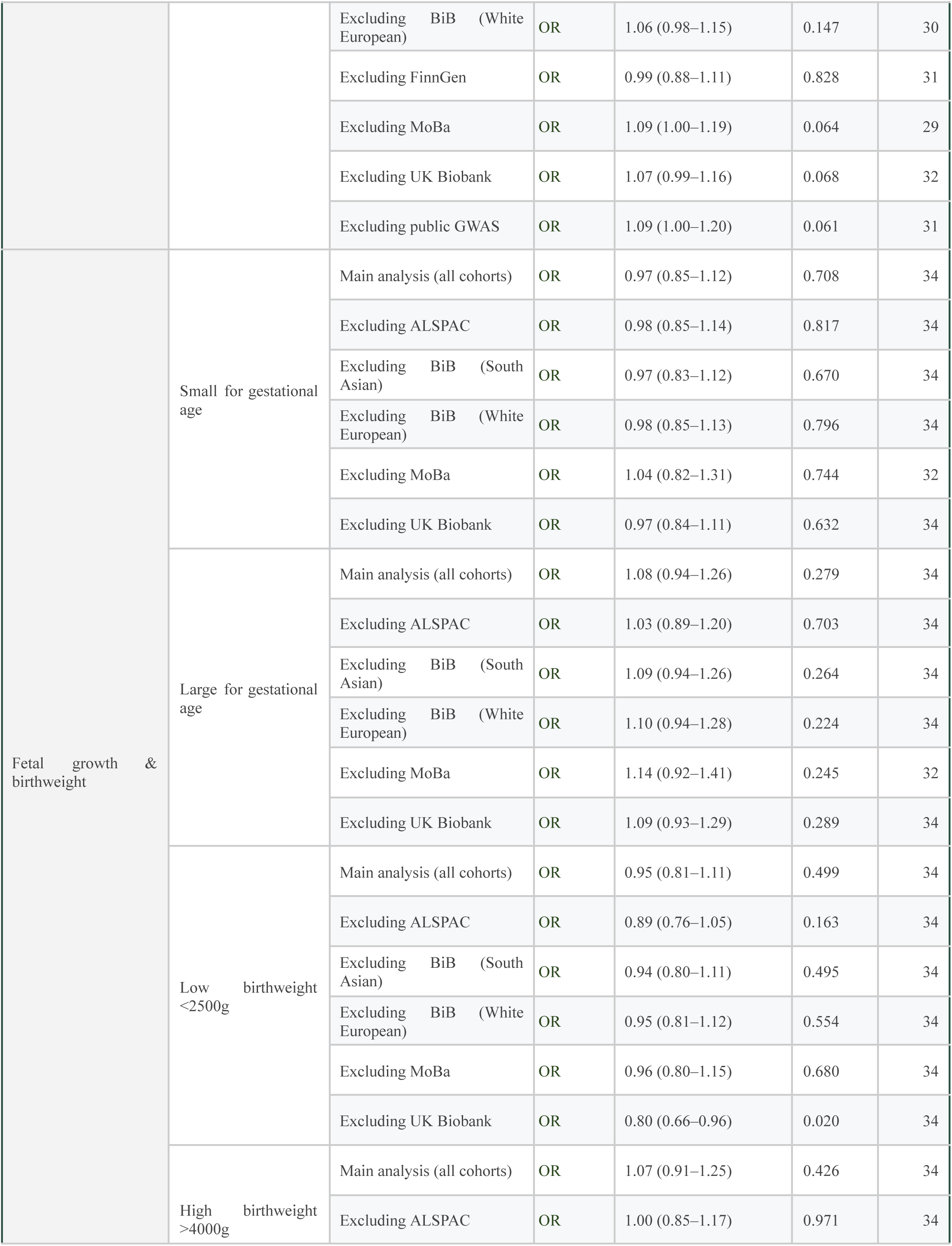
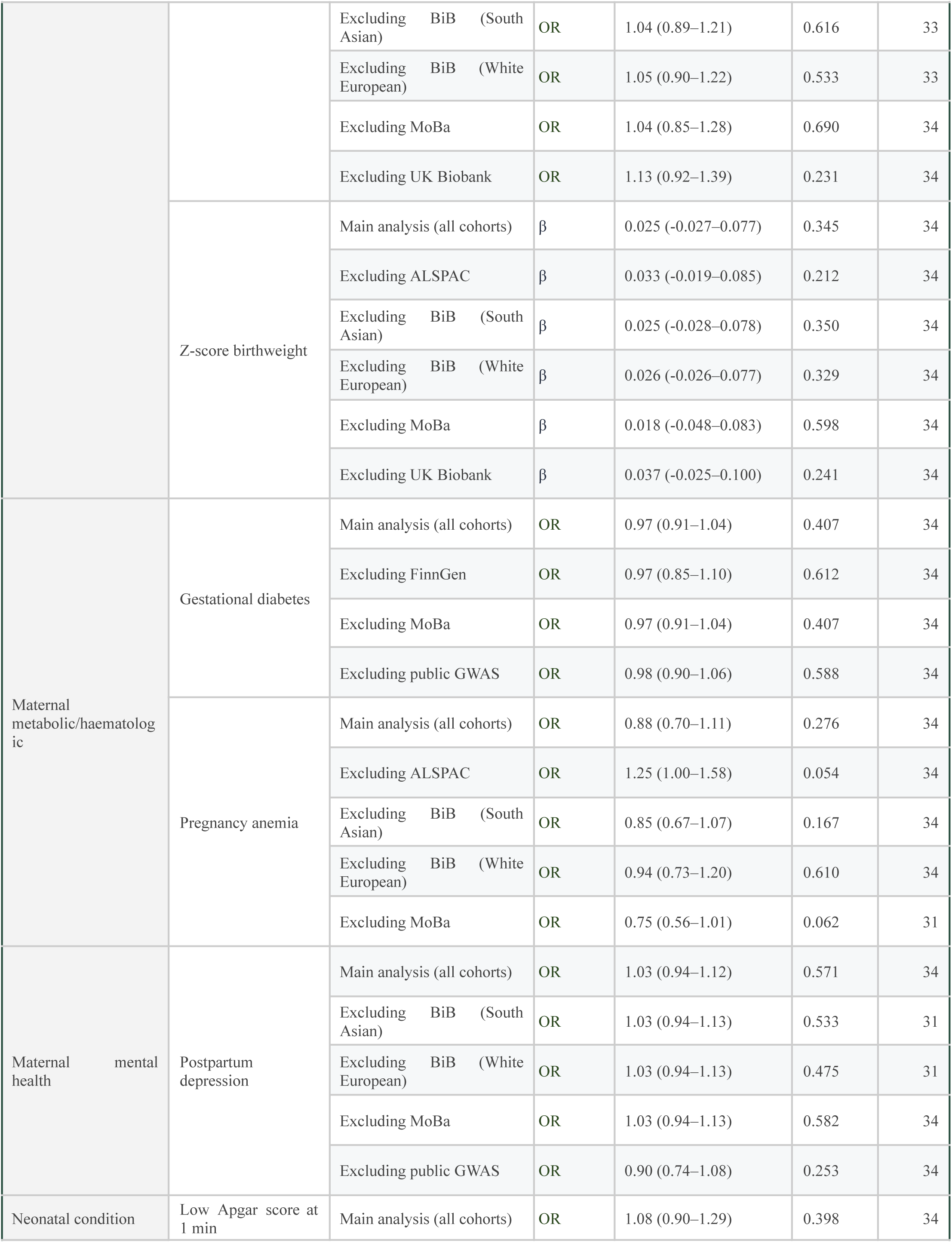
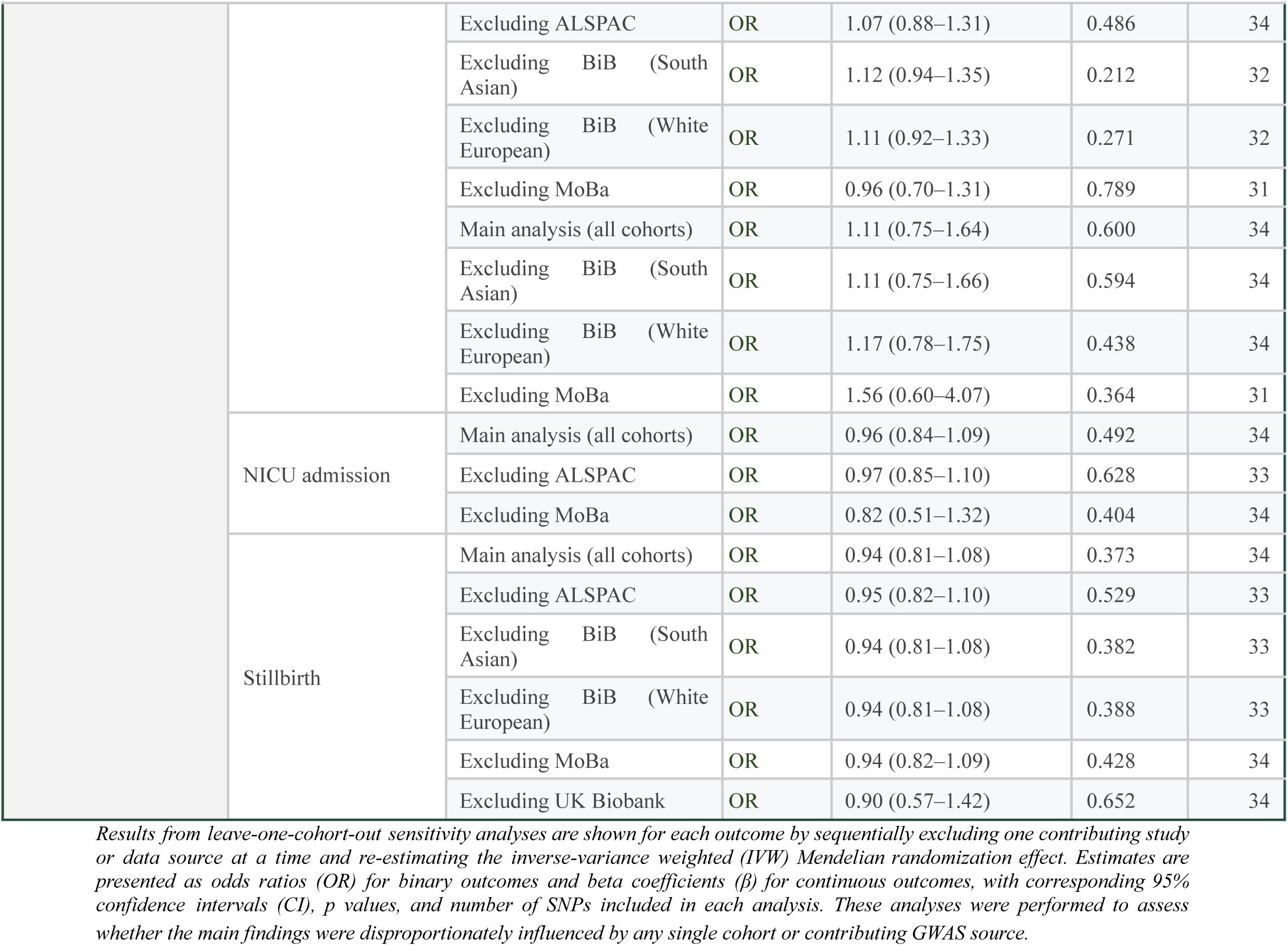
Leave-one-cohort-out sensitivity analysis: IVW estimates across all outcomes with each contributing study sequentially excluded

## SUPPLEMENTARY FIGURES

**Supplementary Figure S1.**
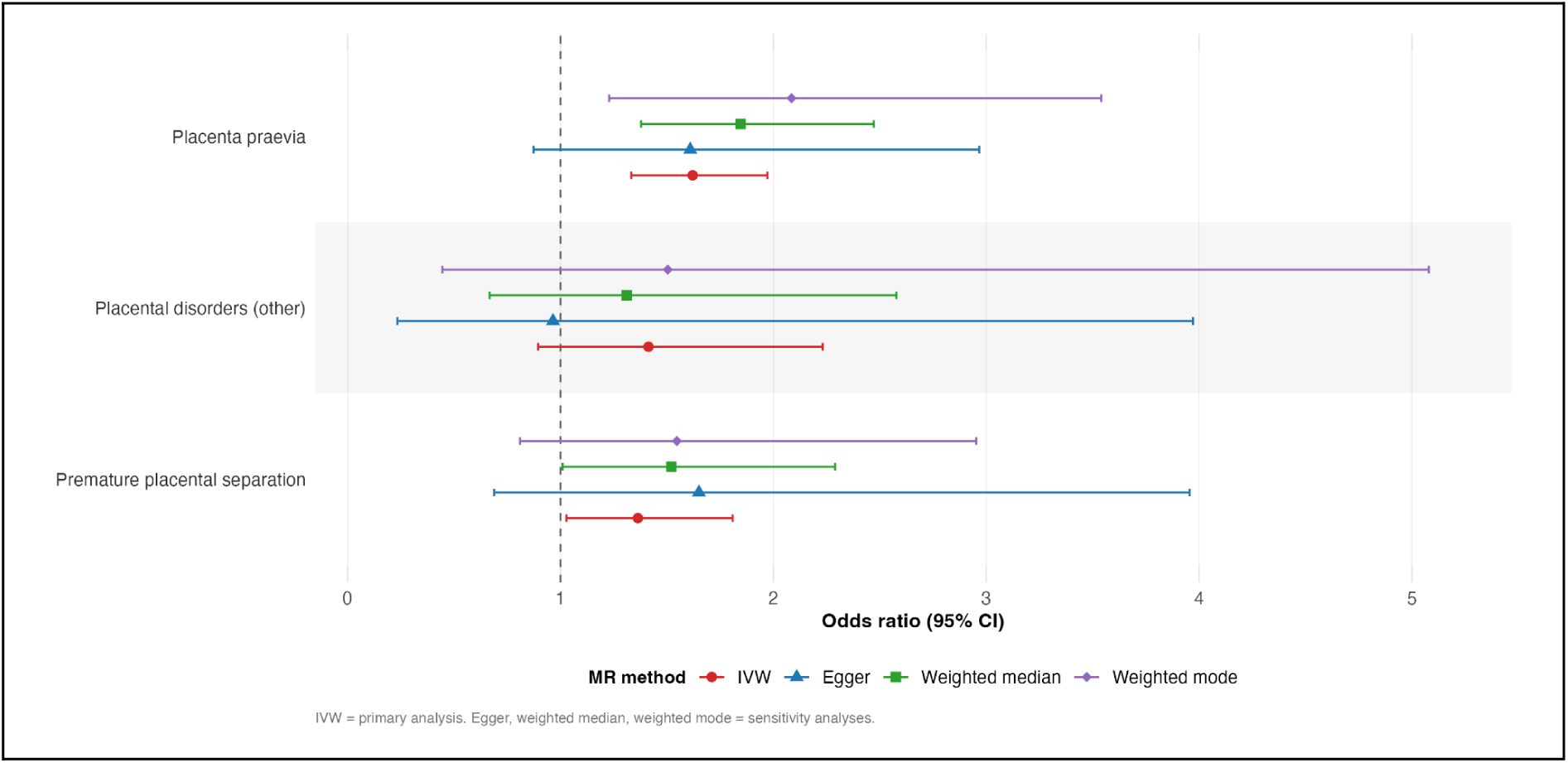
Placental disorders domain across MR methods. *Forest plot showing causal effect estimates of genetic liability to endometriosis on placental outcomes using different Mendelian randomization methods. Results are presented as odds ratios (OR) with 95% confidence intervals for inverse-variance weighted (IVW), MR-Egger, weighted median, and weighted mode approaches. The vertical dashed line represents the null value (OR = 1)*.

**Supplementary Figure S2.**
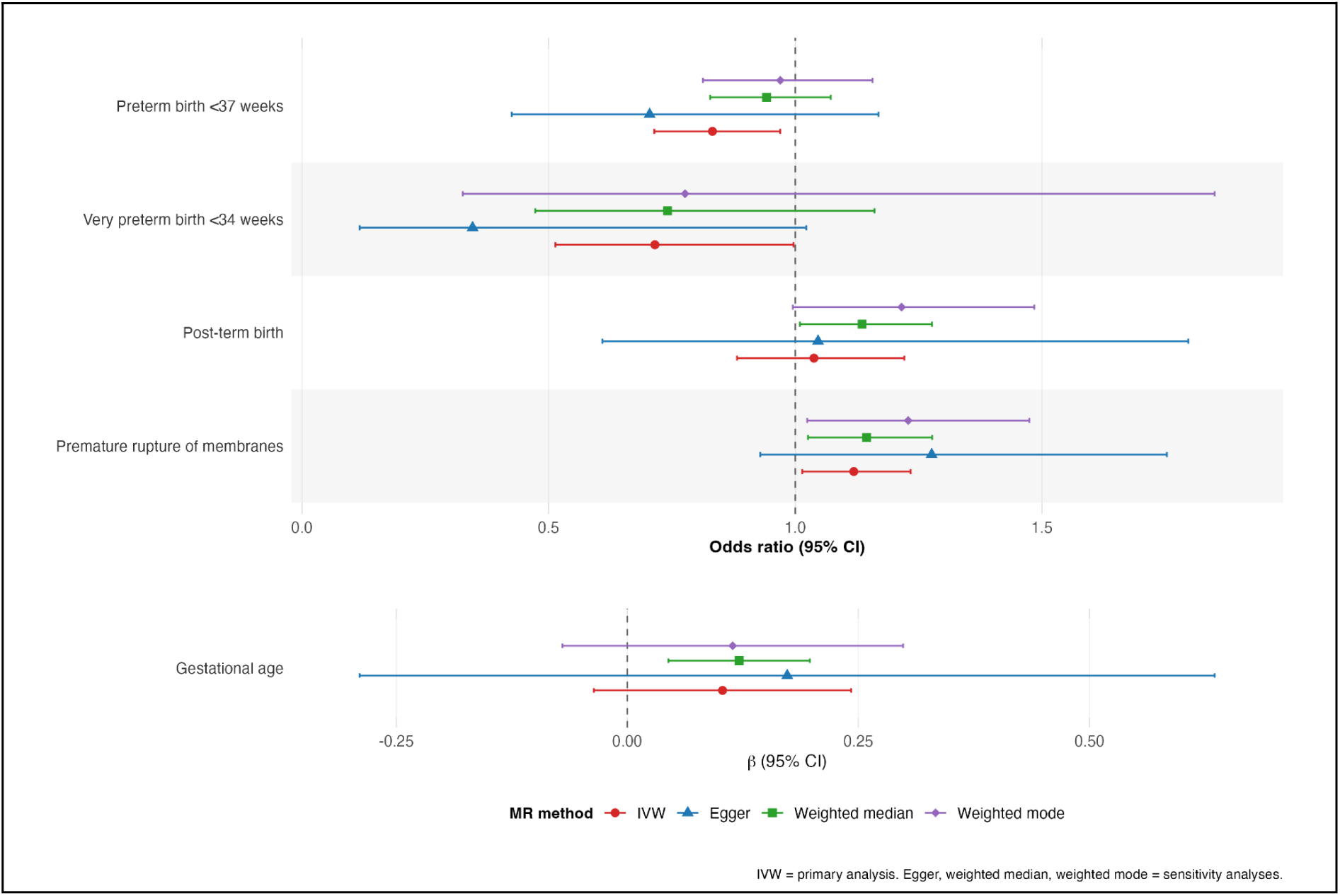
Pregnancy timing domain across MR methods. *Forest plot showing causal effect estimates of genetic liability to endometriosis on pregnancy timing outcomes using inverse-variance weighted (IVW), MR-Egger, weighted median, and weighted mode methods. Results are presented as odds ratios (OR) for binary outcomes and beta (β) coefficients for gestational age, with 95% confidence intervals. The vertical dashed line indicates the null value (OR = 1 for binary outcomes; β = 0 for continuous outcomes)*.

**Supplementary Figure S3.**
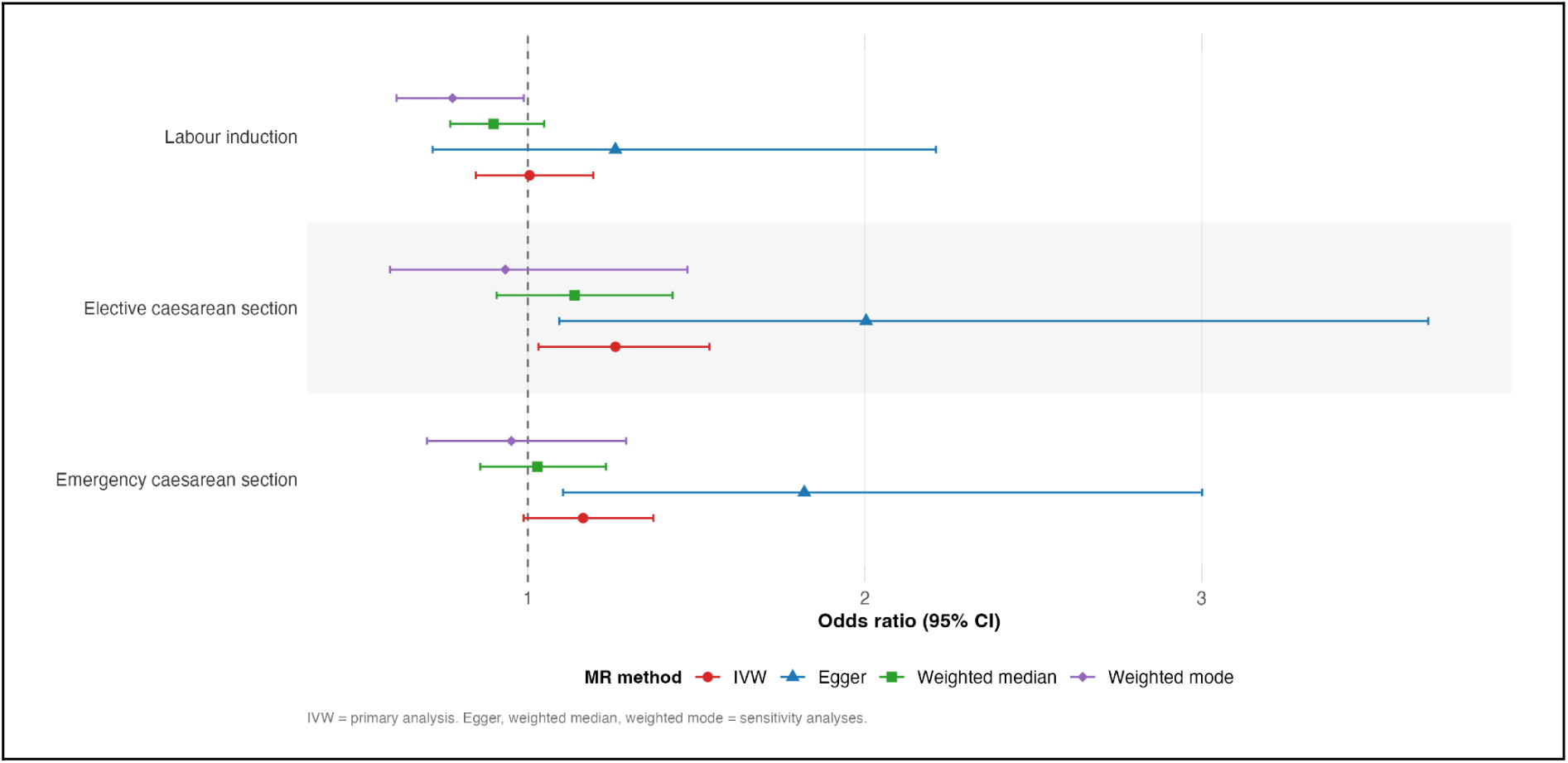
Labour and delivery complications domain across MR methods. *Forest plot showing causal effect estimates of genetic liability to endometriosis on labour and delivery outcomes using inverse-variance weighted (IVW), MR-Egger, weighted median, and weighted mode methods. Results are presented as odds ratios (OR) with 95% confidence intervals. The vertical dashed line represents the null value (OR = 1)*.

**Supplementary Figure S4.**
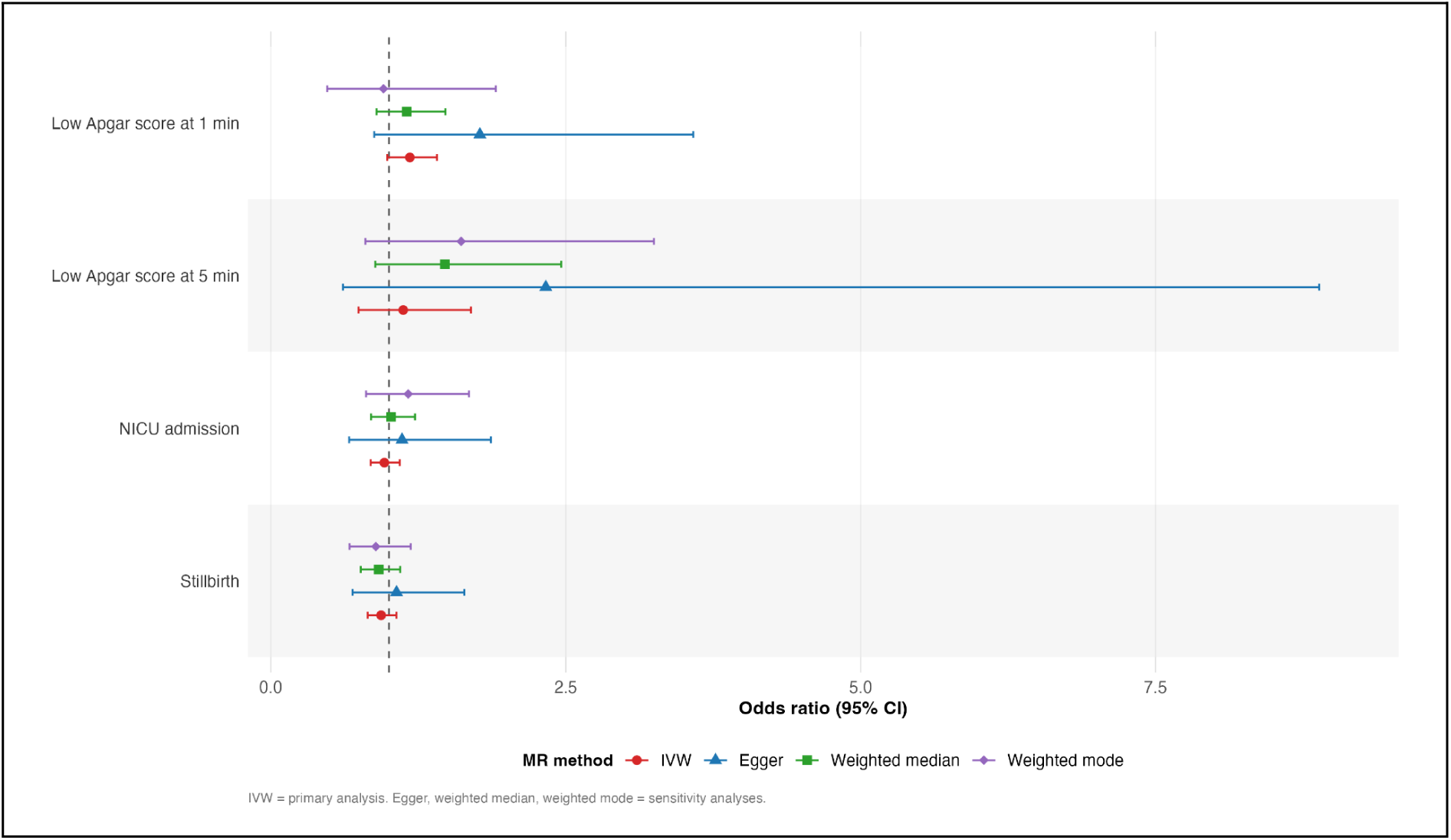
Neonatal outcomes at birth domain across MR methods. *Forest plot showing causal effect estimates of genetic liability to endometriosis on neonatal outcomes using inverse-variance weighted (IVW), MR-Egger, weighted median, and weighted mode methods. Results are presented as odds ratios (OR) with 95% confidence intervals. The vertical dashed line represents the null value (OR = 1)*.

**Supplementary Figure S5.**
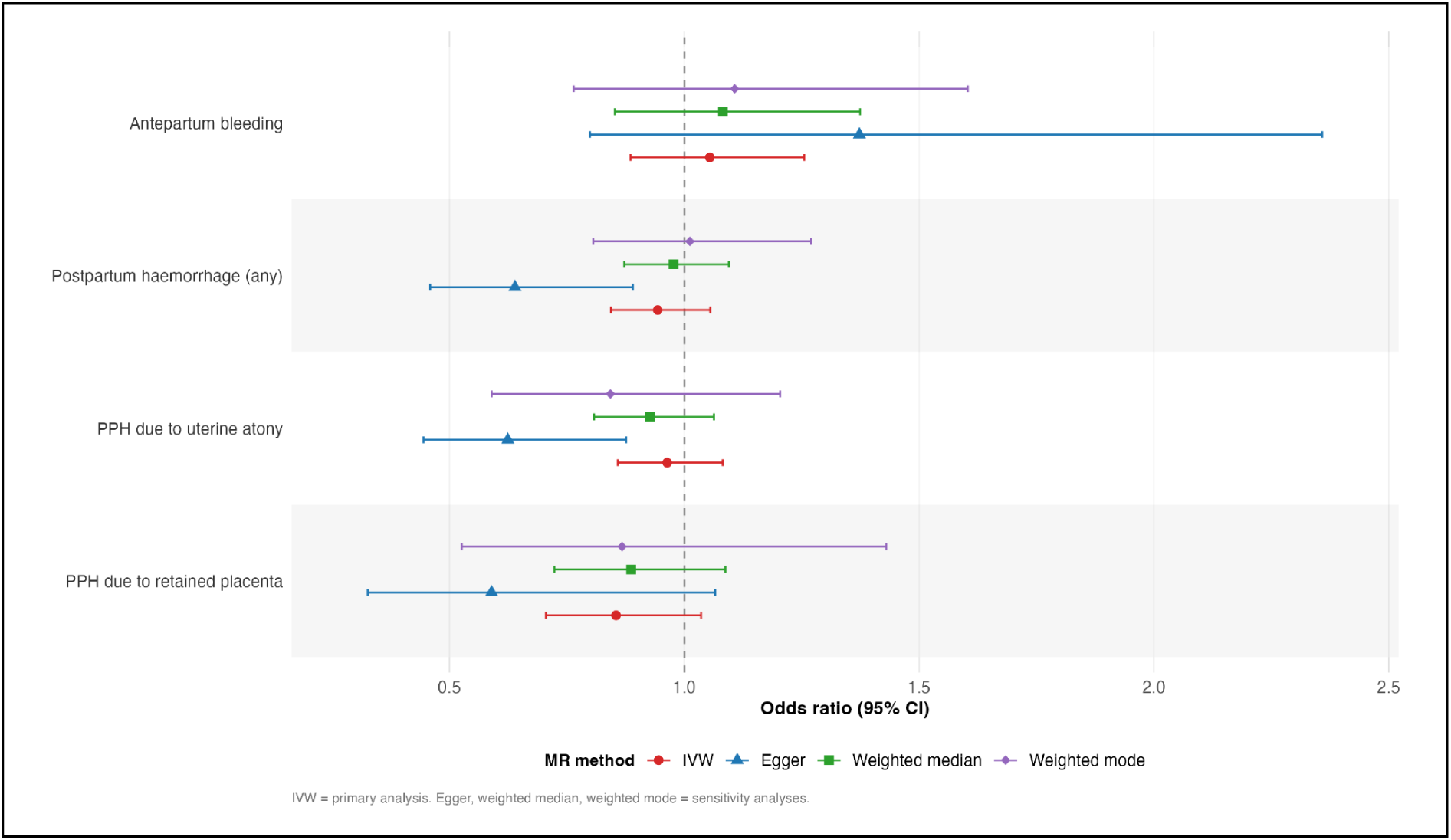
Bleeding and hemorrhage domain across MR methods. *Forest plot showing causal effect estimates of genetic liability to endometriosis on bleeding and haemorrhage outcomes using inverse-variance weighted (IVW), MR-Egger, weighted median, and weighted mode methods. Results are presented as odds ratios (OR) with 95% confidence intervals. The vertical dashed line represents the null value (OR = 1)*.

**Supplementary Figure S6.**
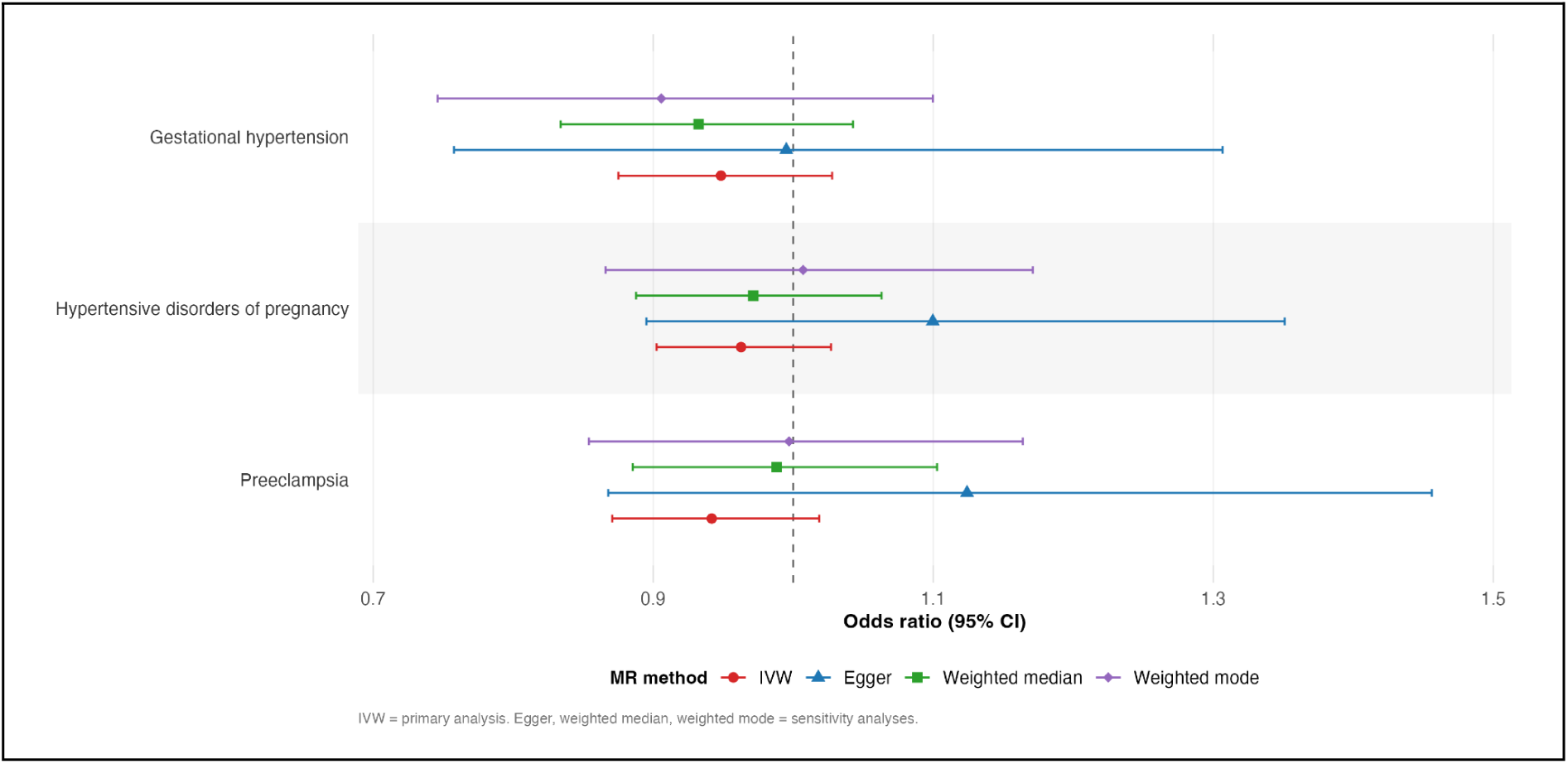
Hypertensive disorders of pregnancy domain across MR methods. *Forest plot showing causal effect estimates of genetic liability to endometriosis on hypertensive disorders of pregnancy outcomes using inverse-variance weighted (IVW), MR-Egger, weighted median, and weighted mode methods. Results are presented as odds ratios (OR) with 95% confidence intervals. The vertical dashed line represents the null value (OR = 1)*.

**Supplementary Figure S7.**
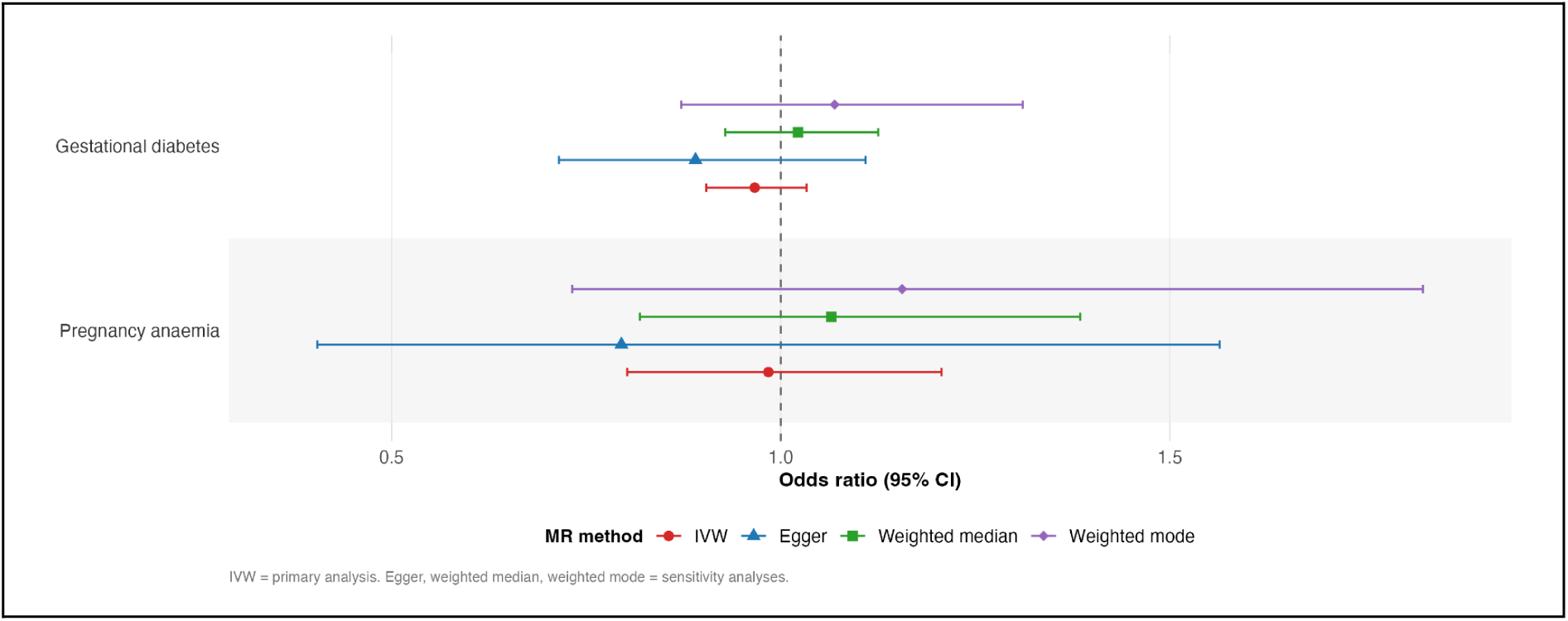
Maternal metabolic/haematologic complications domain across MR methods. *Forest plot showing causal effect estimates of genetic liability to endometriosis on maternal metabolic and haematologic outcomes using inverse-variance weighted (IVW), MR-Egger, weighted median, and weighted mode methods. Results are presented as odds ratios (OR) with 95% confidence intervals. The vertical dashed line represents the null value (OR = 1)*.

**Supplementary Figure S8.**
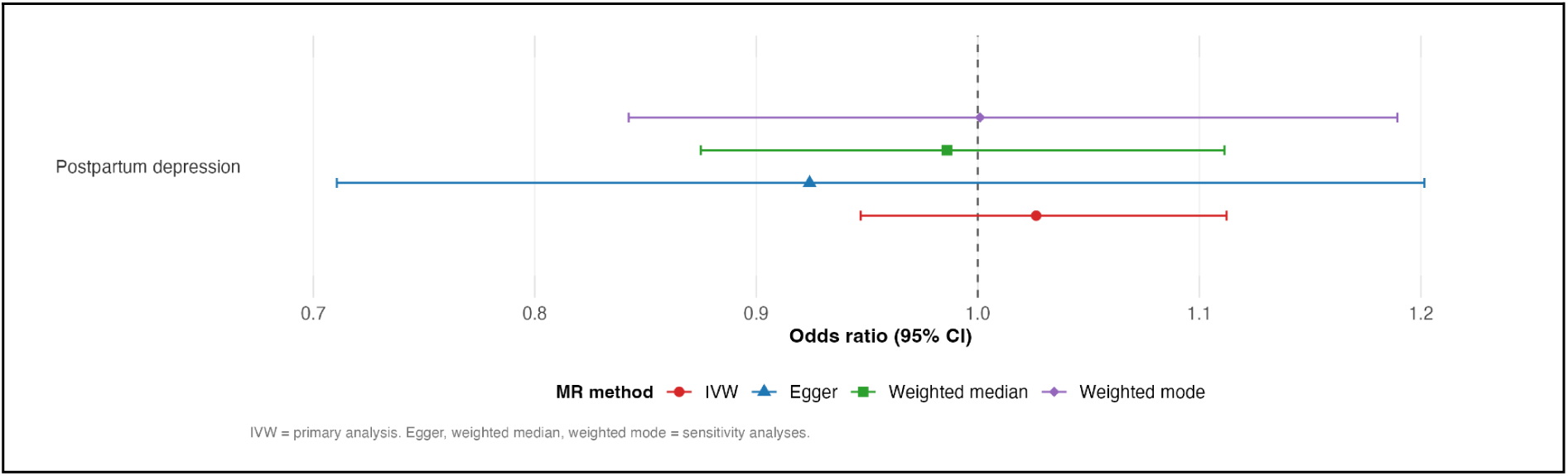
Maternal mental health domain across MR methods. *Forest plot showing causal effect estimates of genetic liability to endometriosis on maternal mental health outcomes using inverse-variance weighted (IVW), MR-Egger, weighted median, and weighted mode methods. Results are presented as odds ratios (OR) with 95% confidence intervals. The vertical dashed line represents the null value (OR = 1)*.

**Supplementary Figure S9.**
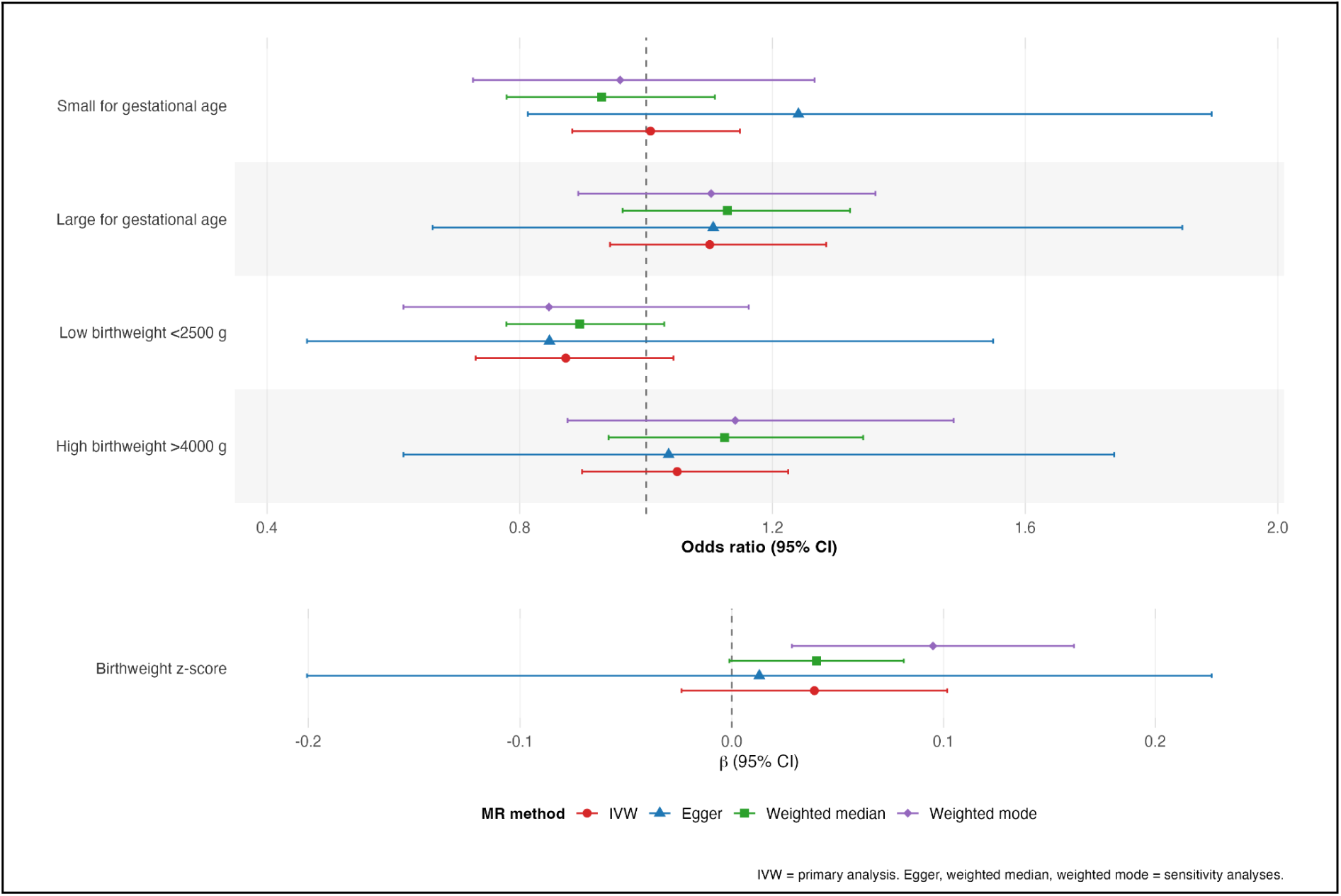
Fetal growth and birthweight domain across MR methods. *Forest plot showing causal effect estimates of genetic liability to endometriosis on fetal growth and birthweight outcomes using inverse-variance weighted (IVW), MR-Egger, weighted median, and weighted mode methods. Results are presented as odds ratios (OR) for binary outcomes and beta (β) coefficients for birthweight z-score, with 95% confidence intervals. The vertical dashed line indicates the null value (OR = 1 for binary outcomes; β = 0 for continuous outcomes)*.

**Supplementary Figure S10.**
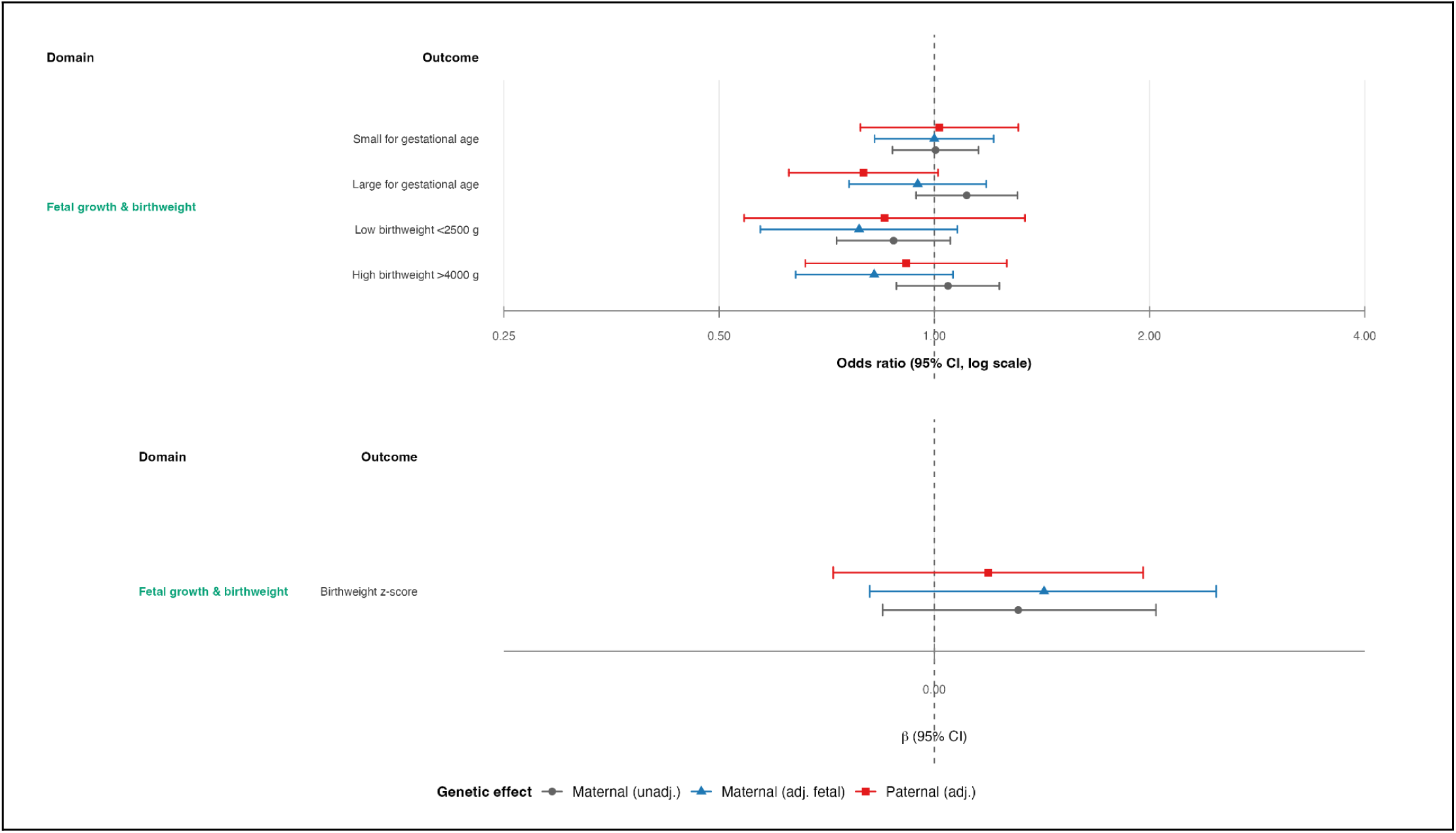
Trio-based Mendelian randomization estimates for birthweight-related outcomes: maternal, fetal, and paternal genetic effects. *Forest plot showing causal effect estimates of genetic liability to endometriosis on birthweight-related outcomes using trio-based Mendelian randomization analyses. Estimates are presented separately for maternal (unadjusted), maternal (adjusted for fetal genotype), and paternal genetic effects. Results are shown as odds ratios (OR) with 95% confidence intervals for binary outcomes and as beta (β) coefficients with 95% confidence intervals for continuous outcomes. The vertical dashed line represents the null value (OR = 1 for binary outcomes; β = 0 for continuous outcomes)*.

**Supplementary Figures S11-16**

*Leave-one-out analyses showing inverse-variance weighted (IVW) estimates for each outcome after sequential exclusion of individual single nucleotide polymorphisms (SNPs). Each point represents the causal estimate obtained after removing one SNP at a time, with horizontal lines indicating 95% confidence intervals*.

**Supplementary Figure S11.**
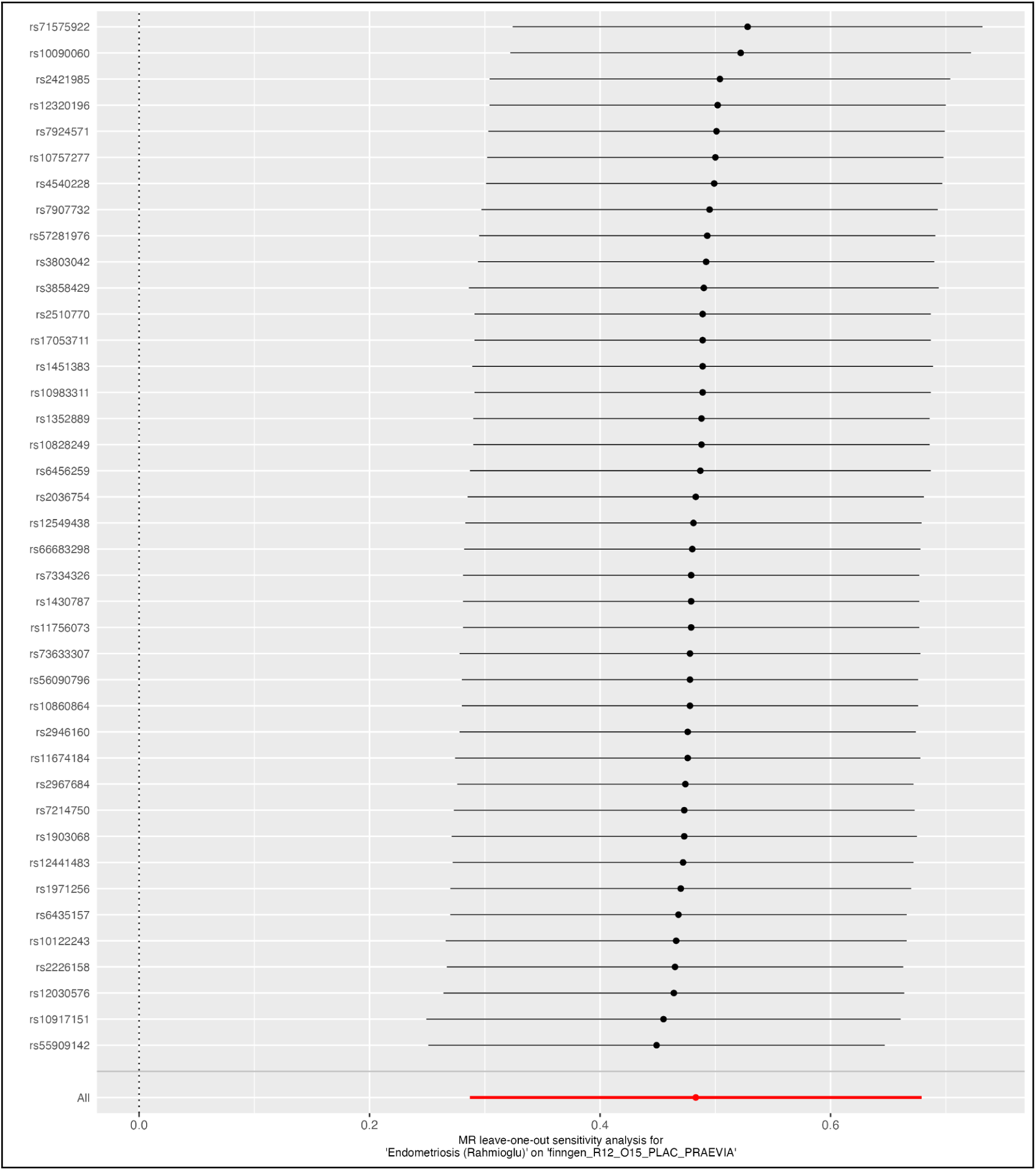
Leave-one-out sensitivity analysis of the effect of endometriosis on placenta praevia, showing the influence of individual SNPs on the IVW estimate.

**Supplementary Figure S12.**
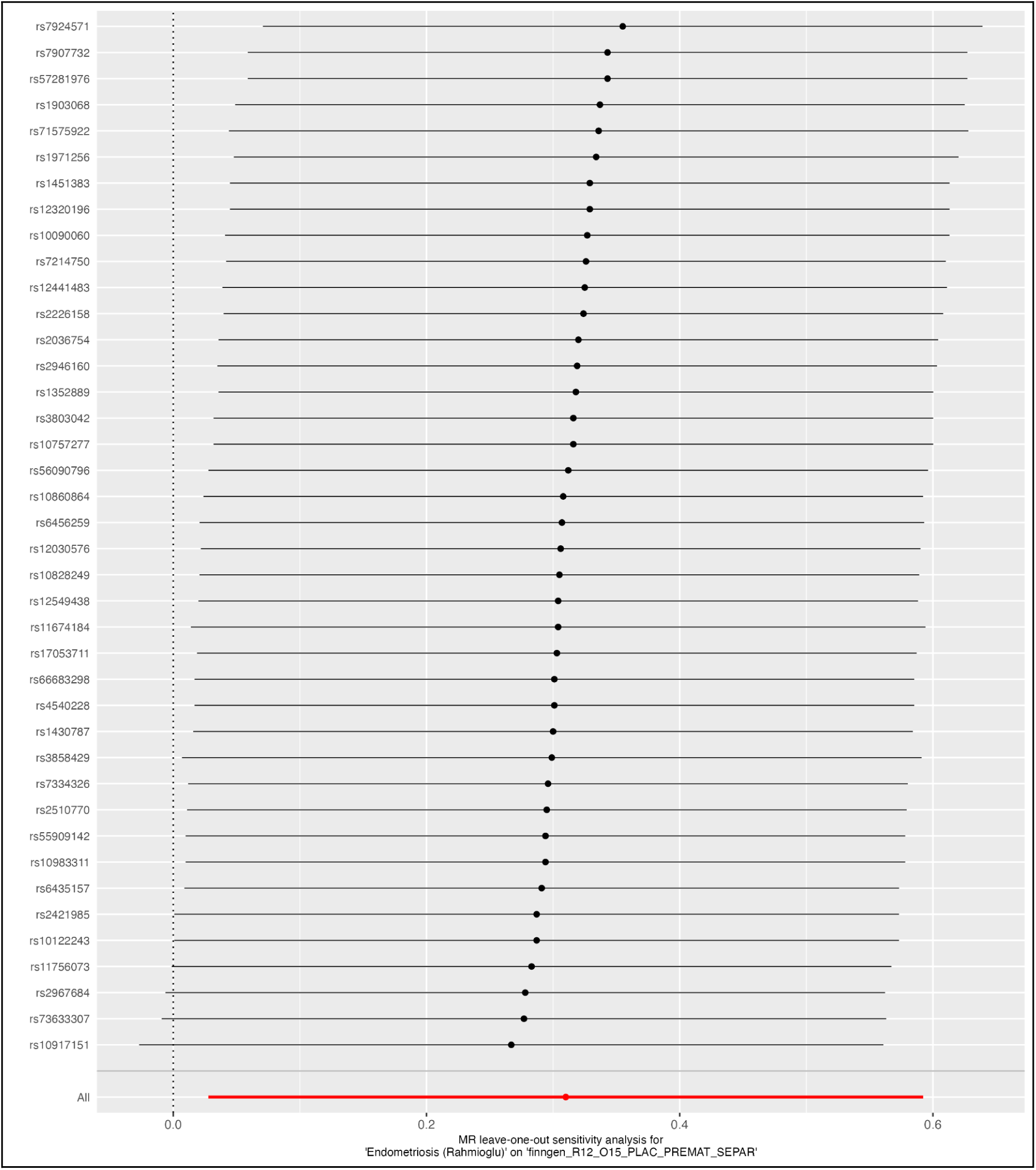
Leave-one-out sensitivity analysis of the effect of endometriosis on premature placental separation, showing the influence of individual SNPs on the IVW estimate.

**Supplementary Figure S13.**
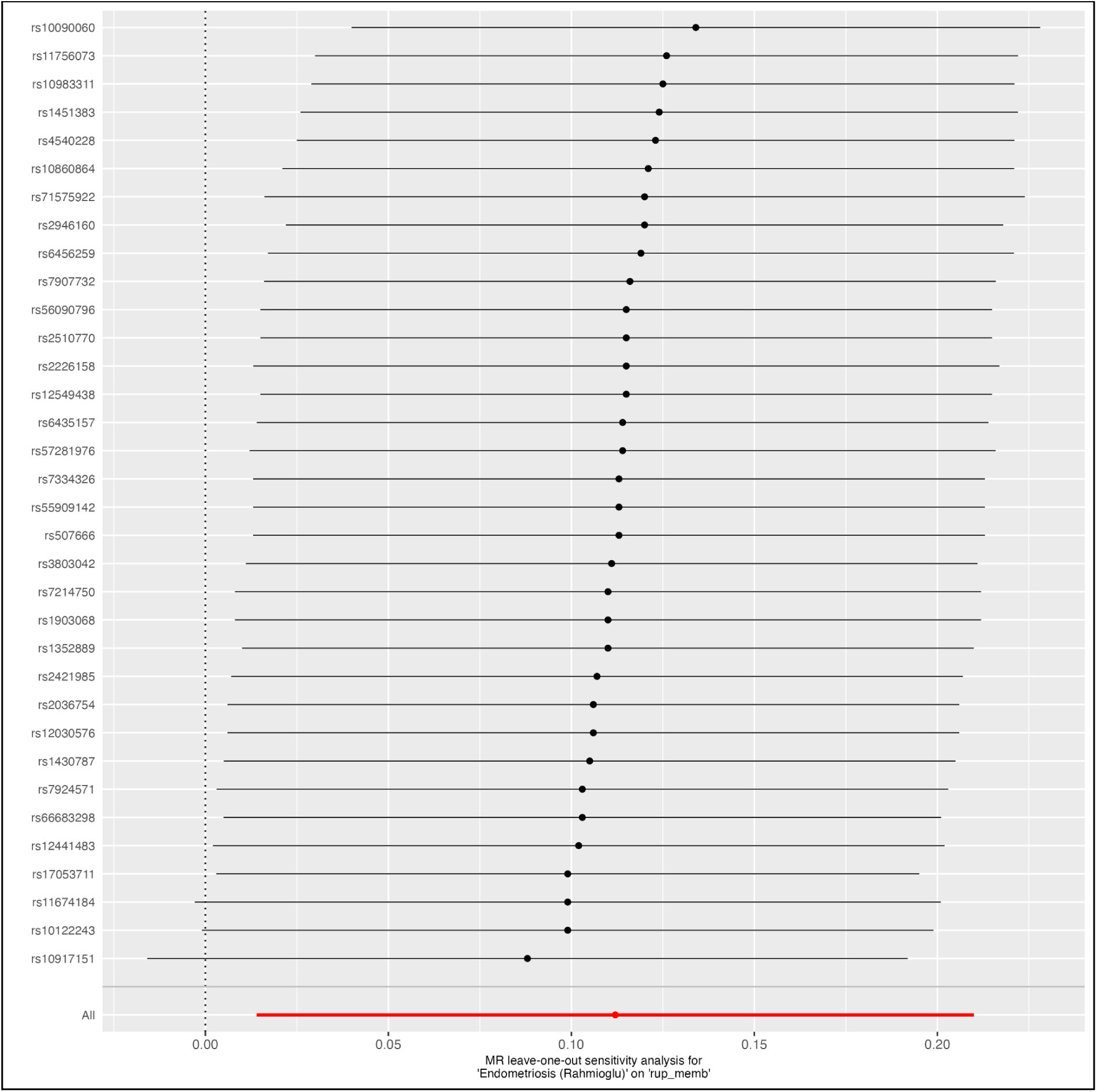
Leave-one-out sensitivity analysis of the effect of endometriosis on premature rupture of membranes, showing the influence of individual SNPs on the IVW estimate.

**Supplementary Figure S14.**
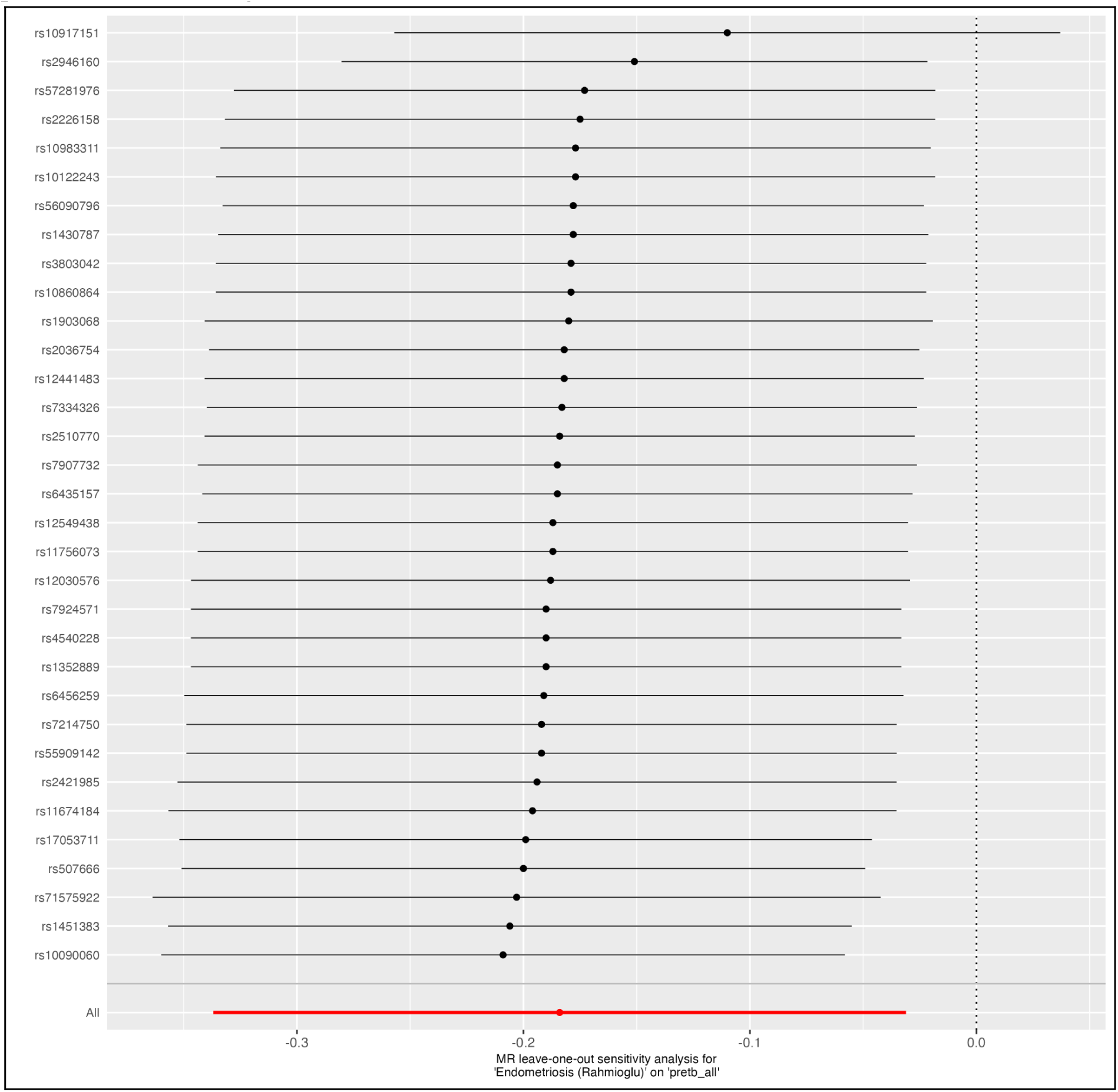
Leave-one-out sensitivity analysis of the effect of endometriosis on preterm birth, showing the influence of individual SNPs on the IVW estimate.

**Supplementary Figure S15.**
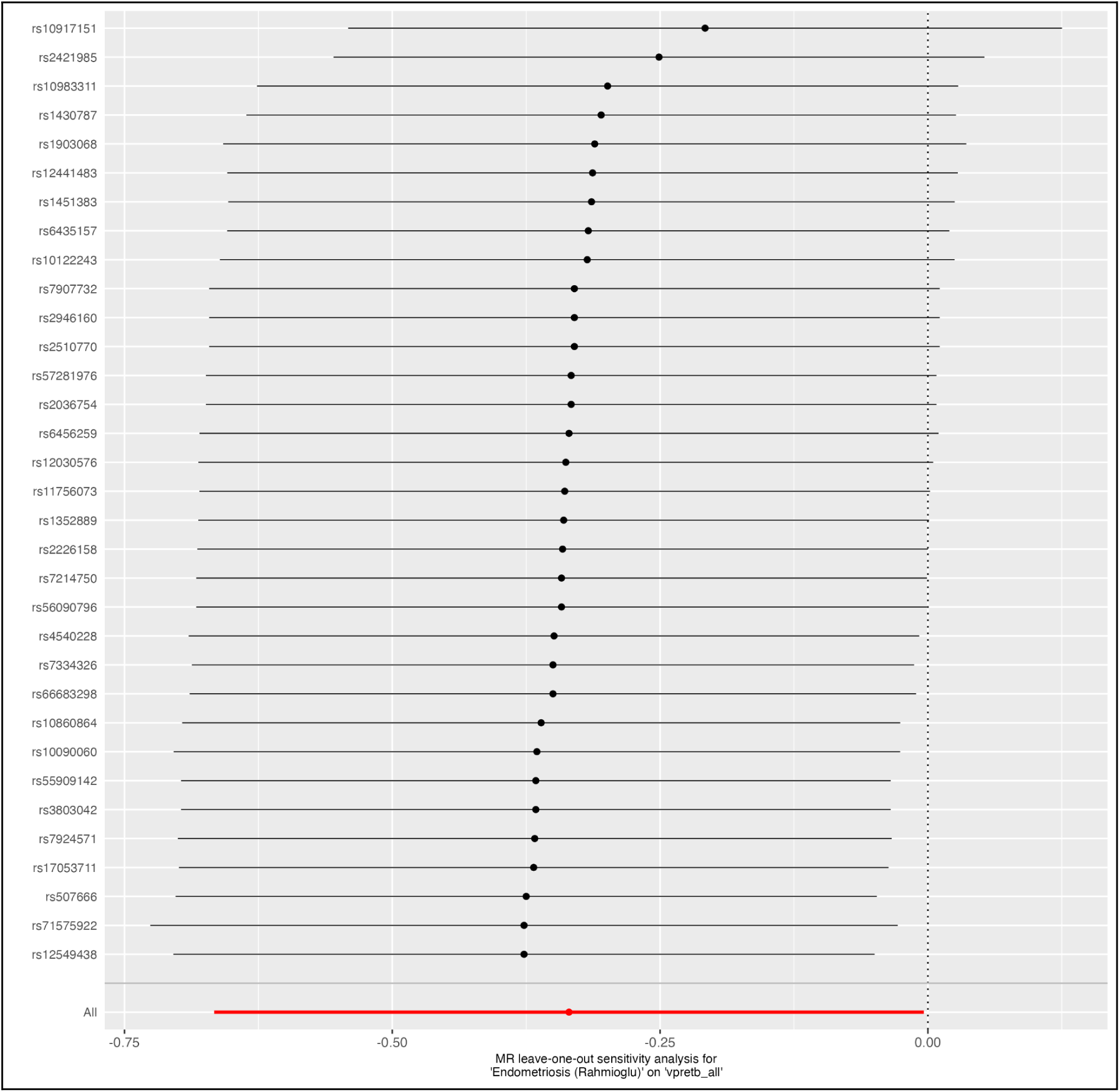
Leave-one-out sensitivity analysis of the effect of endometriosis on very preterm birth, showing the influence of individual SNPs on the IVW estimate.

**Supplementary Figure S16.**
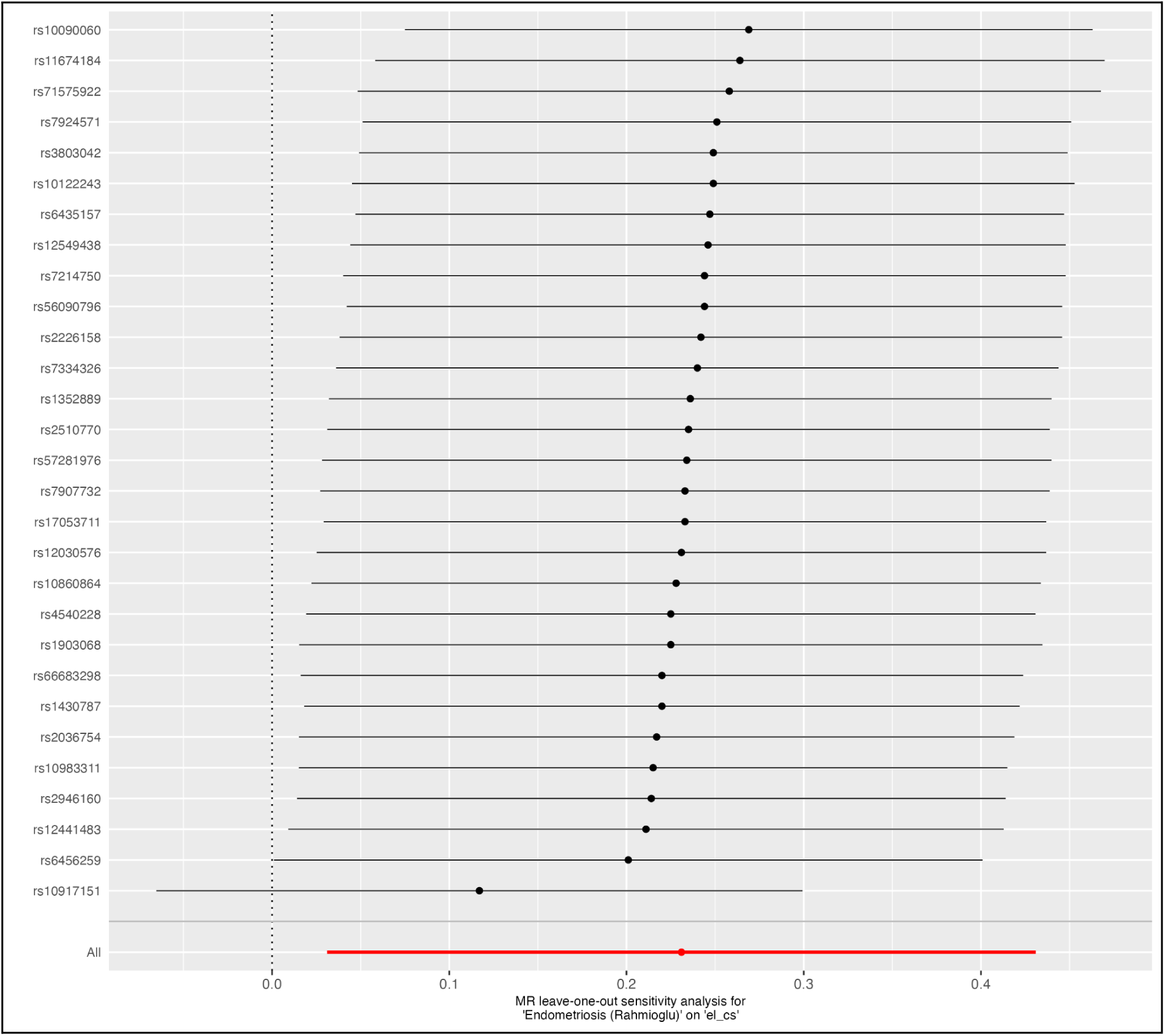
Leave-one-out sensitivity analysis of the effect of endometriosis on elective caesarean section, showing the influence of individual SNPs on the IVW estimate.

