## Supplementary methods for "Genetic liability to endometriosis and pregnancy outcomes: a two-sample Mendelian randomization study with maternal–fetal effect decomposition"

### 1. Exposure Data: Genetic Liability to Endometriosis

#### 1.1. GWAS Source and Variant-Level Quality Control

Genetic instruments for endometriosis were derived from the largest published genome-wide association study (GWAS) meta-analysis of clinically confirmed endometriosis by Rahmioglu et al. (Nature Genetics 2023) (1). This meta-analysis included approximately 60,674 surgically and/or clinically diagnosed cases and 701,926 female controls, predominantly of European ancestry, contributed by the International Endometriosis Genetics Consortium, 23andMe, FinnGen and other cohorts. We imported the European-ancestry summary statistics from the meta-analysis of all cohorts including 23andMe, and restricted all subsequent analyses to these data to maintain ancestry compatibility with the outcome GWAS. We extracted all variants (9,259 SNPs) reaching conventional genome-wide significance for endometriosis ($P < 5 \times 10^{-8}$) from "*Supp33.Top10K-SNPs*", together with their effect and non-effect alleles, effect sizes (beta), standard errors (SE), effect allele frequencies (EAF), and sample size (N). These variants were used to proxy genetic liability to endometriosis. Genome-wide SNP-level summary statistics were extracted from Supplementary Table 33 ("Supp33.Top10K-SNPs") of Rahmioglu et al., which reports an effective sample size of approximately 206,000 individuals per SNP. This effective sample size, reflected in the reported standard errors, was used for all downstream calculations of instrument strength (F-statistics and $R^2$), rather than the overall GWAS sample size. The extracted dataset includes predominantly European-ancestry cohorts and a very small proportion of East Asian participants (<5%). As shown in Supplementary Figure 5 of the original study, effect estimates are highly concordant across ancestries, and the inclusion of East Asian samples had negligible impact on genome-wide associations. To maintain ancestry compatibility with MR-PREG outcome GWAS (which are entirely European), all downstream analyses used effect estimates restricted to European-ancestry populations whenever available. Genome-wide SNP-level summary statistics report an effective sample size of approximately 206,000 individuals per SNP. This effective sample size, reflected in the reported standard errors, was used for all downstream calculations of instrument strength (F-statistics and $R^2$), rather than the overall GWAS sample size. Missing RSIDs were obtained by mapping coordinates using the 1000 Genomes Project Phase 3 European reference panel (2). When multiple rsIDs were present at a given position, we retained the allele-specific identifier consistent with the effect allele reported in the GWAS. Variants with ambiguous mapping or unresolved alleles were excluded at this stage; however, in practice none met these exclusion criteria and all 9,259 genome-wide significant variants were successfully mapped to unique rsIDs.

#### 1.2. Linkage Disequilibrium Clumping

To ensure independence between instruments, we performed linkage disequilibrium (LD) clumping using PLINK 1.9 with 1000 Genomes Phase 3 Europeans as the reference population. We applied an $r^2$ threshold of 0.001 within a 10,000 kb physical window, using $5 \times 10^{-8}$ as the primary P-value threshold and 1 as the secondary threshold. After clumping, 41 independent single nucleotide polymorphisms (SNPs) were retained as instruments for genetic liability to endometriosis.

#### 1.3. Instrument Strength Evaluation

Instrument strength was systematically evaluated. For each SNP, we calculated a simple F-statistic as:

$$F\_simple = (beta / SE)^2$$

where *beta* is the regression coefficient for the SNP–endometriosis association and *SE* is its standard error.

We then estimated the variance explained in endometriosis liability (R^2) using the standard formula (3):

$$R^2 = [\ 2 \times EAF \times (1 - EAF) \times beta^2\ ] / [\ 2 \times EAF \times (1 - EAF) \times beta^2 + SE^2 \times N\ ]$$

where *EAF* denotes the effect allele frequency and *N* the sample size of the exposure GWAS.

From this, we derived an exact per-SNP F-statistic:

$$F\_exact = R^2 \times (N - 2) / (1 - R^2)$$

Summing R^2 across all independent SNPs (assuming negligible LD after clumping) gave a total variance explained of approximately 5.6%. The mean per-SNP F-statistic was approximately 279, substantially above the conventional threshold of 10, indicating a very low risk of weak instrument bias. Full instrument characteristics (rsID, genomic position, alleles, beta, SE, EAF, R^2, F) are provided in Supplementary Table S2.

#### 1.4. Interpretation of Liability in MR Analyses of Categorical Exposures

Endometriosis is a binary clinical diagnosis arising from an underlying continuous liability. In liability-threshold models, genetic variants influence the probability of crossing the diagnostic threshold rather than the binary state itself.

Accordingly, all MR estimates in this study should be interpreted as the effect of a unit increase in genetically predicted liability to endometriosis, rather than as direct causal contrasts between women with and without clinically diagnosed disease.

This conceptual distinction is fundamental for MR analyses of categorical exposures, because genetic instruments act on the latent liability scale. As emphasized by Howe et al. and Burgess et al., MR estimates for binary phenotypes primarily capture the causal effect of underlying liability—which may influence outcomes both through and independently of progression to clinical disease—rather than the effect of the observed exposure category alone (4,5).

This does not invalidate causal interpretation but clarifies that MR-derived effect sizes are not equivalent to risk differences or odds ratios comparing diagnosed versus non-diagnosed individuals in observational epidemiology.

### 2. Outcome Data

We evaluated 30 maternal and perinatal outcomes. These outcomes were chosen based on prior epidemiological evidence linking endometriosis with obstetric complications, clinical relevance for maternal–fetal health, and the availability of adequately powered GWAS in European-ancestry populations. We drew outcome summary statistics from three sources: the Mendelian Randomization in Pregnancy (MR-PREG) collaboration (6), FinnGen release 12 (7), and a large postpartum haemorrhage (PPH) meta-analysis (8).

#### 2.1. MR-PREG Collaboration

MR-PREG is an international consortium that harmonises and meta-analyses GWAS of pregnancy and perinatal outcomes across several large European birth cohorts, including ALSPAC, Born in Bradford, MoBa, UK Biobank, FinnGen and multiple publicly available GWASs, following a standardised pipeline described in detail in McBride et al. (2025) (6). Briefly, each cohort applied standard sample-level quality control (removal of individuals with low call rate (<95-98%), sex discordance, outlying heterozygosity, ancestry outliers and/or cryptic relatedness) and restricted analyses to participants of European genetic ancestry. Variant-level quality control included removal of variants with low call rate (<95-99%), minor allele frequency of <0.5% or <1% or minor allele count <3, violation of Hardy–Weinberg equilibrium (P<1.0e-6) and low imputation quality (<0.4), as described in McBride et al. (6). Association analyses were performed using logistic and linear regressions for binary and continuous traits, respectively, with appropriate adjustment for age, ancestry principal components, batch effects and/or cohort-specific covariates, as well as Firth correction for rare binary outcomes. After excluding studies that overlapped with public GWAS meta-analyses or that contributed fewer than 50 cases for a given outcome, cohort-level results were meta-analysed separately for maternal and offspring genetic effects using fixed-effects inverse-variance weighting. Mutually adjusted maternal and fetal genetic effects at each variant were estimated using a weighted linear model (WLM), which provides maternal genetic effects conditional on offspring genotype (9). Paternal genetic estimates were incorporated as a negative control where available.

From MR-PREG we used 23 outcomes provided through the consortium's meta-analysed GWAS, including gestational age, gestational diabetes, gestational hypertension, hypertensive disorders of pregnancy, high and low birthweight, small-for-gestational-age, birthweight z-score, large-for-gestational-age, preterm birth (<37 weeks), very preterm birth (<34 weeks), post-term birth (>42 weeks), labour induction, premature rupture of membranes, Apgar score <7 at 1 and 5 minutes, NICU admission, pregnancy anaemia, preeclampsia, elective and emergency caesarean section, stillbirth, and postpartum depression. For each outcome GWAS we extracted SNP-level beta, SE, effect allele, allele frequency and sample size directly from the published summary statistics.

#### 2.2. FinnGen Release 12

To complement these outcomes with more detailed placental phenotypes, we used FinnGen release 12 (7). We extracted placenta praevia, placental disorders, and premature placental separation, defined using registry-based ICD-10 codes (O44, O43, and O45, respectively). The FinnGen GWAS follow a standardised QC and analysis framework described in Kurki et al. (2023), including sample-level quality control (i.e. removal of individuals with low call rate (<95%), sex discordance, outlying heterozygosity, non-Finnish ancestry) and variant-level quality control (i.e. removal of variants with low call rate (<98%), minor allele count <3 and violation of Hardy–Weinberg equilibrium (P<1.0e-6). Association analyses were performed using logistic regressions with adjustment for age, ancestry principal components and batch effects.

#### 2.3. Westergaard et al. (Bleeding Outcomes)

Finally, bleeding-related outcomes were obtained from the GWAS meta-analysis by Westergaard et al. (2024) (8), which aggregates data from multiple Nordic biobanks and UK Biobank. We included antepartum bleeding, postpartum haemorrhage (PPH) overall, PPH due to uterine atony and PPH due to retained placenta as defined in that study. Reported odds ratios were converted to log-odds (beta = log(OR)); when standard errors were not available, they were derived from the reported P-values using SE = |beta| / z, where z is the standard normal quantile corresponding to P/2.

#### 2.4. Clinical Domain Organization

Overall, the 30 outcomes can be grouped into nine clinical domains: placental disorders (placenta praevia, placental disorders, premature placental separation); hypertensive disorders of pregnancy (hypertensive disorders of pregnancy (overall), gestational hypertension, preeclampsia); pregnancy timing (gestational age, preterm birth, very preterm birth, post-term birth); fetal growth and birthweight (high birthweight, low birthweight, small-for-gestational-age, birthweight z-score, large-for-gestational-age); labour and delivery complications (labour induction, premature rupture of membranes, elective and emergency caesarean); bleeding and haemorrhage (antepartum bleeding, PPH overall, PPH due to atony, PPH due to retained placenta); maternal metabolic/haematologic complications (gestational diabetes, pregnancy anaemia); maternal mental health (postpartum depression); and neonatal condition at birth (low Apgar scores at 1 and 5 minutes, NICU admission, stillbirth). Detailed definitions, case/control counts and contributing cohorts are presented in Table 2 and Supplementary Table S1.

### 3. Data Harmonisation

#### 3.1. Harmonisation Procedure

Harmonisation of endometriosis and outcome GWAS was carried out using the TwoSampleMR package (version 0.5.6) with additional manual checks. For each outcome, we first matched the 41 endometriosis instrument SNPs to outcome SNPs by rsID. We then aligned alleles so that the effect allele was identical in exposure and outcome datasets, flipping the sign of the outcome beta and adjusting allele frequencies where necessary. Palindromic SNPs with intermediate allele frequencies (A/T or C/G variants with minor allele frequency around 0.42), for which strand alignment cannot be reliably inferred using allele frequency information, were flagged during harmonisation using the harmonise_data function with action = 2. In the present study, all 7 palindromic SNPs had allele frequencies sufficiently distant from 0.5 to allow reliable strand inference, and none were flagged for removal. Variants flagged as unsuitable for MR analysis (mr_keep = FALSE) were subsequently excluded. While all 41 original instruments survived global harmonisation QC, the final number of SNPs available per outcome ranged from 29 to 40, with some SNPs unavailable in specific outcome GWAS datasets due to missing summary statistics or outcome-specific QC failures. When exposure SNPs were absent from a given outcome GWAS, proxy SNP substitution was not used in the primary analysis. A targeted proxy search was explored during sensitivity checks, but only one suitable proxy variant was identified and its inclusion did not materially alter the estimates; to ensure consistency across outcomes and avoid introducing proxy-specific assumptions, the main analyses were based on direct rsID matching only.. The exact SNP count per outcome is reported in Table 2, alongside exposure and outcome sample sizes.

### 4. Sample Overlap

Partial sample overlap is possible between the exposure GWAS and some outcome datasets, particularly UK Biobank and FinnGen, which contributed to both the endometriosis GWAS by Rahmioglu et al. (1) and several outcome GWASs from MR-PREG and FinnGen. Sample overlap in two-sample MR can bias estimates toward the confounded observational association when instrument strength is weak (10). However, given the strong instrument strength in this study (mean F-statistic ≈ 279, total variance explained ≈ 5.6%), any bias arising from sample overlap is expected to be minimal. Simulation studies have shown that with F-statistics substantially above 10, the impact of sample overlap on bias is negligible (10,11). Additionally, the use of summary-level data precludes precise quantification of individual-level overlap. To further assess robustness, we conducted cohort-level leave-one-out analyses for MR-PREG outcomes (see Section 5.6), which confirmed that no single cohort—including those with potential overlap—disproportionately influenced the results.

### 5. Statistical Analyses

#### 5.1. **Primary MR Analysis (IVW)** (10)

For each outcome, causal effects were primarily estimated using the inverse-variance weighted (IVW) method (10), which combines SNP-specific Wald ratios using inverse-variance weighting. For analyses including three or more SNPs, estimates corresponded to the multiplicative random-effects IVW model implemented in the TwoSampleMR package, allowing for heterogeneity between instrument-specific estimates.

For binary outcomes, causal estimates were exponentiated to obtain odds ratios (OR = exp(beta)) with 95% confidence intervals, whereas for continuous outcomes (gestational age, birthweight z-score) beta coefficients and corresponding 95% confidence intervals were reported.

#### 5.2. **MR-Egger Regression** (12)

We performed MR-Egger regression to evaluate the robustness of IVW estimates to potential violations of MR assumptions, particularly horizontal pleiotropy. First, we performed MR-Egger regression, modelling beta_Yi as:

$$\beta_{Yi} = \text{intercept} + \beta_{MR\_Egger} \times \beta_{Xi} + \text{error}$$

The slope beta_MR_Egger provides a pleiotropy-adjusted causal estimate under the Instrument Strength Independent of Direct Effect (InSIDE) assumption, while the intercept tests for directional pleiotropy: a non-zero intercept suggests that instruments have, on average, direct effects on the outcome independent of endometriosis liability.

#### 5.3. **Weighted Median and Mode Estimators** (13)

We estimated causal effects using the weighted median estimator, which remains consistent if at least 50% of the total instrument weight is contributed by valid instruments. This estimator is based on the median of SNP-specific ratio estimates (beta_Yi / beta_Xi), weighted by the inverse variance of beta_Yi. Where informative, we additionally computed weighted mode estimates, which assume that the most common (modal) causal estimate across SNPs arises from valid instruments.

#### 5.4. **MR-PRESSO** (14)

We applied the MR-PRESSO (Pleiotropy RESidual Sum and Outlier) method (MRPRESSO package v1.0.0). For each outcome, the global test examined whether the pattern of residuals was compatible with no horizontal pleiotropy. When the global test was significant, MR-PRESSO identified outlier SNPs and recomputed outlier-corrected IVW estimates after excluding them. The distortion test was used to assess whether removal of outliers significantly altered the causal estimate. We report global test P-values, outlier SNPs where present and corrected effect estimates.

#### 5.5. **Heterogeneity Assessment** (15)

Third, we quantified heterogeneity among SNP-specific causal estimates using Cochran's Q statistic. For each SNP we calculated its ratio estimate beta_i = beta_Yi / beta_Xi and weight w_i = beta_Xi^2 / var_Yi, and then computed:

$$Q = \text{sum}(\, w_i \times (\beta_i - \beta_{IVW})^2 \,)$$

A large Q statistic with a low P-value indicates heterogeneity and may suggest pleiotropy or violation of model assumptions.

#### 5.6. Leave-One-Out Analyses (SNP and Cohort Level) (16,17)

Finally, we performed single-SNP MR and leave-one-SNP-out analyses, standard diagnostic procedures in MR to assess the influence of individual variants on causal estimates (16,17). Single-SNP estimates examine the contribution of each instrument individually, while leave-one-out analyses re-run IVW MR after removing each SNP in turn to evaluate whether any single variant unduly drives results. In addition, we conducted cohort-level leave-one-out analyses for MR-PREG outcomes, following standard meta-analytic influence diagnostics (18), by re-generating SNP-outcome meta-analysis summary statistics after sequential exclusions of each contributing cohort, followed by repeating IVW MR analyses. Figures summarising SNP-level leave-one-out analyses are presented in Supplementary Figures S11–S16, while results of cohort-level leave-one-out analyses are reported in Supplementary Table S5.

#### 5.7. Consistency Assessment

Sensitivity analyses (MR-Egger, weighted median, weighted mode) were used as diagnostic procedures to assess robustness to pleiotropy, rather than as independent hypothesis tests; accordingly, FDR correction was not applied to these estimates. Consistency between sensitivity and primary IVW results was formally evaluated based on two criteria: (i) concordance in effect direction, and (ii) overlap of the sensitivity method's 95% confidence interval with the IVW point estimate. We did not require sensitivity analyses to reach nominal significance, as these estimators have substantially lower statistical power than IVW due to relaxed assumptions (MR-Egger) or reduced efficiency (weighted median/mode). Therefore, wider confidence intervals and loss of statistical significance in sensitivity analyses were expected and not interpreted as evidence of inconsistency, provided effect directions were concordant and confidence intervals overlapped with the IVW estimate. Using these criteria, 17/30 (57%) outcomes showed full consistency across all sensitivity methods, and all nominally significant IVW associations demonstrated full consistency (Supplementary Table S3).

In addition to statistical significance, interpretation of causal estimates incorporated consideration of effect magnitude, direction, and precision. Magnitude was evaluated in relation to clinical and biological plausibility, recognising that very small effect sizes may have limited clinical relevance despite statistical significance. Directional consistency across MR methods was considered supportive of robustness. Precision was assessed based on the width of confidence intervals, with imprecise estimates interpreted cautiously even when nominally significant. This framework was applied to contextualise findings in the presence of correlated outcomes and varying statistical power across analyses.

#### 6. **Maternal, Fetal, and Paternal Genetic Effects** (19)

Some outcomes, such as hypertensive disorders of pregnancy, birthweight and preterm birth, may be influenced by both maternal and fetal genomes (19,20). Where available, we therefore leveraged trio-based genetic effect estimates from MR-PREG to distinguish maternal and fetal pathways.

MR-PREG provides effect estimates from models jointly including maternal, offspring (fetal) and paternal genotypes, derived from mother–father–child trios or extended parent–offspring structures, using regression models of the form

expected outcome = beta_m × G_m + beta_o × G_o + beta_p × G_p + covariates

, where GmG_mGm, GoG_oGo and GpG_pGp represent maternal, offspring and paternal genotypes. Maternal effects (βm\beta_mβm) capture direct maternal genetic influence on the intrauterine environment, fetal effects (βo\beta_oβo) reflect the fetus's own genome acting on fetal growth and development, and paternal effects (βp\beta_pβp) serve as a negative control, as paternal genotype should not directly affect maternal pregnancy complications.

Trio-based genetic associations were available for 21 of the 30 investigated outcomes, namely small-for-gestational-age, large-for-gestational-age, high birthweight (>4000 g), low birthweight (<2500 g), birthweight z-score, gestational age, post-term birth, preterm birth <37 weeks, preeclampsia, gestational hypertension, hypertensive disorders of pregnancy, gestational diabetes, pregnancy anemia, labour induction, emergency caesarean section, elective caesarean section, low Apgar score at 1 minute, low Apgar score at 5 minutes, neonatal intensive care unit admission, postpartum depression and premature rupture of membranes.

For each outcome, the primary Mendelian randomization analyses used maternal genetic effects on the pregnancy outcome, using estimates adjusted for fetal genotype when available. The same maternal-effect SNP–endometriosis associations were used as the exposure dataset in all analyses. Additional analyses were performed using fetal genetic effects on the outcomes and paternal genetic effects, the latter serving as a negative control to reduce potential collider bias and residual familial confounding. These analyses were considered sensitivity analyses. Thus, while the exposure data remained identical across analyses, the SNP–outcome associations differed according to whether maternal, fetal, or paternal genetic effects were modelled. The inverse-variance weighted (IVW) method was used as the primary causal estimator in each pathway. Additional Mendelian randomization methods (MR-Egger, weighted median, and weighted mode), together with heterogeneity statistics, MR-Egger intercepts, MR-PRESSO global and outlier tests, and leave-one-out analyses, were performed as sensitivity analyses to assess the robustness of the findings. Because not all trio-based SNP associations were available for all outcomes, the number of instruments contributing to each maternal, fetal, and paternal analysis varied accordingly, ranging from 27 to 34 SNPs (details in Supplementary Table S4). Results of these analyses, allowing direct comparison of maternal, fetal and paternal pathways, are reported in Figure 3. Detailed estimates are provided in Supplementary Table S4, and birthweight-specific trio-based analyses are presented in Supplementary Figure S10.

The maternal–fetal genotype correction (WLM) could be applied only to outcomes from the MR-PREG consortium, where maternal, paternal and/or fetal genotype estimates were available. For FinnGen and Westergaard et al. datasets, which provide only maternal GWAS summary statistics, adjusted maternal effects cannot be derived. As a result, effect estimates for these outcomes represent unadjusted maternal effects. MR-PREG outcomes are presented with and without fetal adjustment to allow comparison.

### 7. **Multiple Testing Correction** (21)

Because we evaluated causal estimates for 30 outcomes, we controlled for multiplicity using the false discovery rate (FDR). We applied the Benjamini–Hochberg procedure to the two-sided IVW P-values across all outcomes using R's p.adjust function in R with method = "fdr". The resulting FDR-adjusted P-values (q-values) reflect the expected proportion of false positives among declared significant findings. We considered associations with $q < 0.05$ as statistically significant after FDR correction, and we report both nominal P-values and q-values.

### 8. **Software and Reproducibility**

Statistical analyses were performed in R version 4.3.2 (R Foundation for Statistical Computing). Data harmonisation and Mendelian randomization analyses were conducted using the TwoSampleMR package (version 0.5.6), with pleiotropy and outlier detection performed using MR-PRESSO (version 1.0.0). Data preparation and management used data.table and dplyr, and visualisations were generated using ggplot2. LD clumping was performed using PLINK version 1.9. The complete analysis pipeline, including scripts (01–07) and documentation, is publicly available in the endoMR-PREG repository (https://github.com/jonasvibert/endoMR-PREG), ensuring full reproducibility.

### 9. Data Availability and Ethics

This study relied exclusively on de-identified summary-level GWAS data from previously approved studies and consortia. Endometriosis GWAS summary statistics were obtained from Rahmioglu et al. (Nature Genetics 2023) (1). MR-PREG outcome GWAS were accessed through the MR-PREG collaboration under its data-sharing policies (6). FinnGen summary statistics (release 12) are publicly available through the FinnGen results portal (7), and bleeding outcomes were obtained from Westergaard et al. (Nature Genetics 2024) (8). Because only aggregated summary statistics were used and no individual-level genetic or clinical data were accessed, the present analyses do not constitute human subjects research as defined by 45 CFR 46.102, and no additional institutional review board approval was required. All original contributing studies obtained appropriate ethical approvals and informed consent from participants.

### 10. Funding of contributing cohorts

Funding information for contributing cohorts and GWAS sources is summarised below.

**ALSPAC** The UK Medical Research Council and Wellcome (Grant ref: 217065/Z/19/Z) and the University of Bristol provide core support for ALSPAC. A comprehensive list of grants funding is available on the ALSPAC website (http://www.bristol.ac.uk/alspac/external/documents/grant-acknowledgements.pdf). ALSPAC GWAS data was generated by Sample Logistics and Genotyping Facilities at Wellcome Sanger Institute and LabCorp (Laboratory Corporation of America) using support from 23andMe. This research was funded in part by the Wellcome Trust (Grant ref: 217065/Z/19/Z). For the purpose of Open Access, the author has applied a CC BY public copyright licence to any Author Accepted Manuscript version arising from this submission.

**MoBa** MoBa funding is under Acknowledgements as requested by MoBa publication guidelines.

**Born in Bradford (BiB)** BiB receives core funding from the Wellcome Trust (WT101597MA), a joint grant from the UK Medical and Economic and Social Science Research Councils (MR/N024397/1), British Heart Foundation (CS/16/4/32482), and the National Institute of Health Research under its Applied Research Collaboration for Yorkshire and Humber and Clinical Research Network research delivery support. Further support for genome-wide and multiple 'omics measurements in BiB is from the UK Medical Research Council (G0600705), National Institute of Health Research (NF-SI-0611-10196), US National Institute of Health

(R01DK10324), and the European Research Council under the European Union's Seventh Framework Programme (FP7/2007–2013) / ERC grant agreement no 669545.

**UK Biobank** UK Biobank is funded primarily by the Wellcome Trust and the Medical Research Council (MRC). It is also funded by the Department of Health, British Heart Foundation, Cancer Research UK, Diabetes UK, National Institute for Health Research (NIHR), Scottish Government, Northwest Regional Development Agency, and Welsh Assembly Government.

**Endometriosis GWAS (Rahmioglu et al., Nature Genetics 2023)** Funding details for the endometriosis GWAS meta-analysis are provided in the original publication (Rahmioglu et al., 2023), including contributions from the International Endometriosis Genetics Consortium, 23andMe, FinnGen, and other participating cohorts.

**Postpartum Haemorrhage GWAS (Westergaard et al., Nature Genetics 2024)** Funding details for the GWAS meta-analysis of postpartum haemorrhage are reported in the original publication (Westergaard et al., 2024), which includes contributions from FinnGen, the Danish Blood Donor Study Genomic Consortium, the Estonian Biobank, the Norwegian Mother, Father and Child Cohort Study (MoBa), and other Nordic cohorts. The study received support from the Novo Nordisk Foundation (NNF17OC0027594, NNF14CC0001) and the US National Institutes of Health (R01 HD101669).

**FinnGen (Release 12)** FinnGen is funded by Business Finland (grants HUS 4685/31/2016 and UH 4386/31/2016) and industry partners. Full funding details, including participating biobanks and contributors, are available at: https://finngen.gitbook.io/documentation/
